# Does de-identification of data from wearable Biometric Monitoring Technologies give us a false sense of security? A systematic review

**DOI:** 10.1101/2022.10.04.22280658

**Authors:** Lucy Chikwetu, Yu Miao, Melat K. Woldetensae, Diarra Bell, Daniel M. Goldenholz, Jessilyn Dunn

## Abstract

It remains unknown whether de-identifying wearable biometric monitoring data is sufficient to protect the privacy of individuals in the dataset. This systematic review seeks to shed light on this. We searched Web of Science, IEEE Xplore Digital Library, PubMed, Scopus, and the ACM Digital Library on December 6, 2021 (PROSPERO CRD42022312922). We also performed manual searches in journals of interest until April 12, 2022. Though our search strategy had no language restrictions, all retrieved studies were in English. We included studies demonstrating re-identification, identification, or authentication using data from wearables. Our search returned 17,625 studies, and 72 studies met our inclusion criteria. Our findings demonstrate that substantial re-identification risk exists in data from sensors generally not thought to generate identifiable information, such as the electrocardiogram and electromyogram. In many cases, only a small amount of data (1-300 seconds of recording) is sufficient for re-identification.

## Introduction

Is there a privacy concern when sharing de-identified data from wearable biometric monitoring technologies? This question has never been more pressing. The wearable device market (USD 116·3 billion in 2021) is projected to reach USD 265·4 billion by 2026.^1^ Some wearables have proven medical applications, for example, detecting arrhythmias^2^ or infections.^3^ Generally, data from wearables is persistent^4^ and has the potential to be shared widely to improve the accuracy and generalizability of algorithms. To support such advancements, the National Institutes of Health (NIH) has adopted policies encouraging extensive data sharing practices starting in 2023.^5^ Additionally, many institutions are adopting the Findable Accessible Interoperable and Reusable (FAIR) Guiding Principles for scientific data management and stewardship.^6^ While data sharing provides tremendous benefits, it also opens up many critical questions surrounding privacy risks to patients and study participants that remain unanswered. For example, could machine learning algorithms be applied to public datasets or data shared through third-party data sharing agreements to enable re-identification? Is there an opportunity for data misuse by governments, corporations, or individuals? If so, how significant is this risk, and is there a way to mitigate it?

Here, we define re-identification as the act of determining an individual’s identity from deliberately de-identified/anonymized data. Re-identification often involves re-linking a de-identified/anonymized dataset with a dataset with identifiers to establish users present in both. Merely matching data does not constitute re-identification. Instead, there is a need for identifiers for re-identification to take place. In real-world scenarios, identifiers are not always available; however, unscrupulous entities who want to know more about individuals whose data they already possess may have them (Figure 1). In addition, data breaches^7^ can also lead certain individuals/entities to possess a complete list or subset of identifiers. For this review, we assume motivated individuals gain access to identifiers and build machine learning algorithms to re-link/match biometric signals.

**Figure 1.**
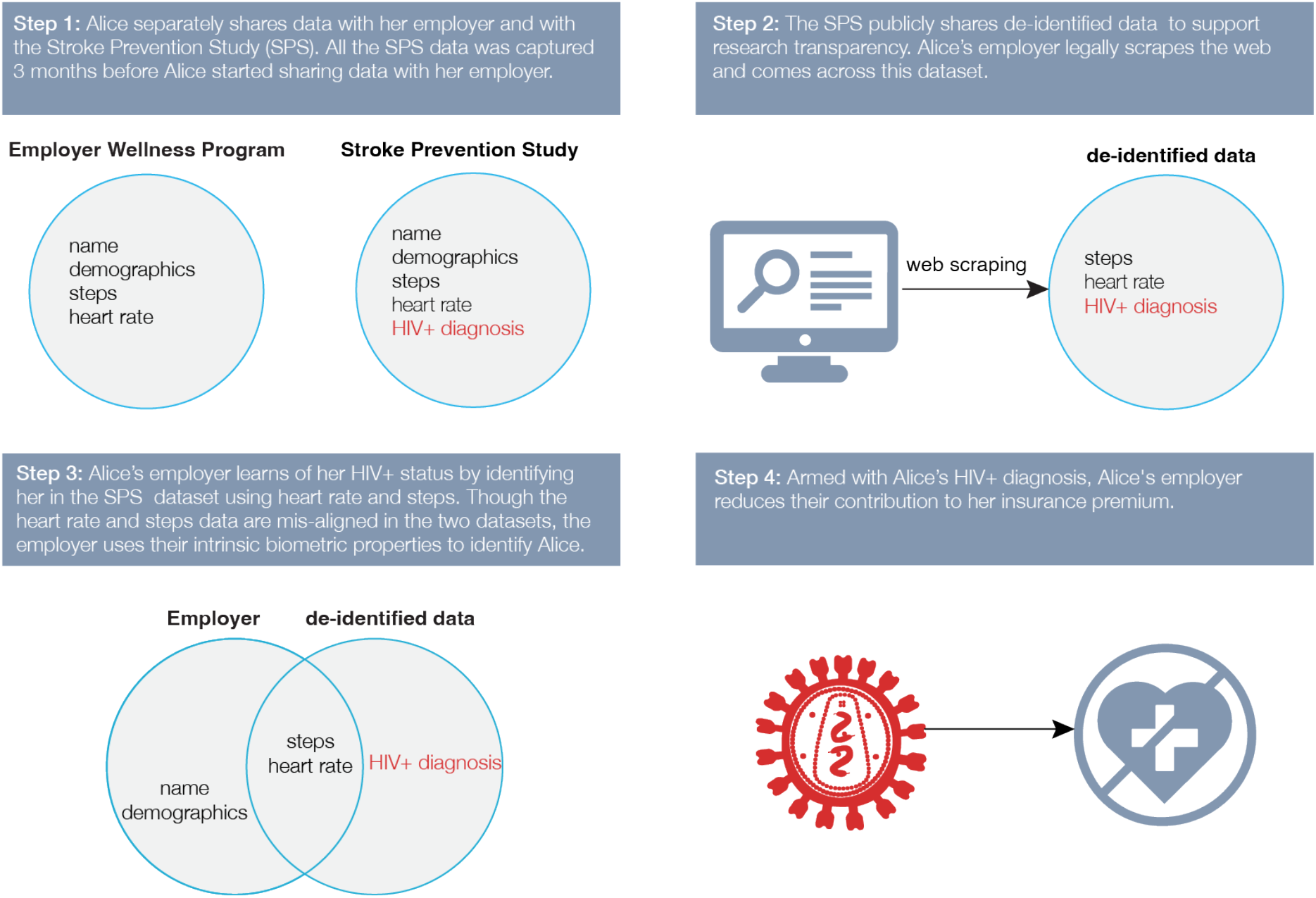
An example scenario with an HIV+ employee who does not wish to share their HIV status with their Employer, but this information is divulged unintentionally through data sharing with the Stroke Prevention Study.

As a result of re-identification, the release of seemingly innocuous data can have unforeseen consequences. One notable example is the re-identification of the Massachusetts Governor from publicly shared and seemingly de-identified state employee health insurance data^8,9^, which led to the passage of the Health Insurance Portability and Accountability Act (HIPAA) in 1996^10^. This example also demonstrates that regulation changes often lag behind real-world re-identification events and their consequences. With biomedical data, the consequences of re-identification may be dire (e.g., Figure 1). Advances in machine learning have made it possible to infer sensitive information about individuals, such as their medical diagnoses^11^, mental health^12^, personality traits^13^, and emotions^14^, thus making it possible to learn information that an individual has not directly shared. Re-identification, therefore, can reveal not only the initially collected data but also such inferences about an individual.

Fundamentally, data from any sensing modality that can create a unique digital identifier (i.e., a “fingerprint”) can potentially be used for biometric identification/authentication (e.g., iris scans and face scans). Any such data may be used to re-identify an individual.^15^

This paper explores open questions surrounding re-identification through an extensive systematic review of available literature. For example, what types of wearable data, how much of that data, and what resolution of such data can enable re-identification are all critical questions that remain unanswered. Our goal is to provide an overview of re-identification risks from wearables often not considered as generating identifiable information.

## Methods

This review follows Preferred Reporting Items for Systematic Reviews and Meta-Analyses (PRISMA)^16^ guidelines. It is registered on PROSPERO (CRD42022312922).

### Information Sources

We searched peer-reviewed literature indexed in Web of Science, IEEE Xplore Digital Library, PubMed, Scopus, and the ACM Digital library on December 6, 2021, with no date limits. We also searched previously identified journals of interest. In April 2022, we identified a recently published review article^17^ exploring some of the topics discussed here; however, our review employed a broader search strategy, found additional sensing modalities, and delved deeper into re-identification. Additionally, our review focuses on the biomedical research community, which is newly charged with public data requirements^5^. We used Covidence software^18^ to conduct this review.

### Search Strategy

This review focused on re-identification using biometric signals from wearables as opposed to other forms of re-identification, such as camera-based re-identification. We exclude GPS-based technologies or biometrics widely used for identification (e.g., iris scans and fingerprints), as these present clear privacy risks.^19^ Although we primarily focus on studies conducted using wearables, to highlight what could be possible, we also report findings from currently uncommon wearables such as the seismocardiogram and the phonocardiogram, even if the measurement modality was not a wearable form factor. The keywords we used for our searches were (*re-id\** OR *reid\** OR *identi\**) and (*biometric\** OR *biosensor\**) together with a bag of words identifying sensors such as *“acceleromet*”, “gyroscope”, “ECG”, “PPG”*, and *“phonocardiogra*”* (appendix p 2-5). Given search functionality restrictions in IEEE Xplore, we decomposed the IEEE Xplore database search into multiple separate searches that met the required guidelines. Our search strategy did not restrict the language in which articles were published; however, all retrieved studies were in English.

### Eligibility Criteria

All included studies were peer-reviewed journal and conference papers published before April 12, 2022. Eligible studies had to demonstrate re-identification, identification, or authentication using biometric signals collected in humans using wearables except in circumstances of rare sensors such as the phonocardiogram, which could include biometric monitoring technologies with non-wearable form factors. Additionally, we only included studies with unimodal sensors since we did not come across any studies where unimodal sensors failed to perform re-identification when used independently yet succeeded when combined with other sensors.

We excluded studies that: used animals, used theoretical models, used video/cameras, employed impractical form factors (such as multiple inertial measurement unit (IMU) sensors attached to five locations ^20^), did not describe sensor placement, had unclear sensor specifications, or did not report standard performance metrics. We also excluded 28 studies with similar findings to other studies by the same authors.

### Screening and Selection

We exported all studies to Covidence^18^, which automatically identified and removed 6,218 duplicates. Two independent reviewers performed title and abstract screening and full-text review, while a third reviewer acted as the adjudicator for resolving inter-rater disagreements. LC ensured quality assurance of the process, and all reviewers resolved any resulting anomalies after adjudication.

### Data Extraction and Synthesis

Two reviewers independently extracted data and performed study quality and publication bias assessments for each study while an adjudicator resolved all conflicts. If any included study referenced other studies meeting our eligibility criteria in any tables in that study’s paper, we also extracted information from those referenced studies. We extracted 18 study characteristics from the included studies (appendix p 5-6) and sensing-modality-specific characteristics: the number of channels/leads (ECG, EEG, and EMG) and the evoked potential stimulus (EEG).

To minimize error, LC reviewed the extracted data for potential discrepancies, and all team members resolved any identified issues. Finally, all graphs were generated in R(v4.0.2) using ggplot2.

## Results

Our search retrieved 17,625 studies (6,218 duplicates), resulting in 11,407 studies to be screened (Figure 2). After title and abstract screening, 1,012 studies advanced to the full-text review. Of these, 65 met the eligibility criteria. We then performed a nested search for additional relevant studies in all of the tables in the 65 studies and uncovered an additional 12 studies. We also removed five studies from the original 65 since they were review articles that referenced other studies we had included. Finally, we extracted data from included studies and subsequently analyzed them (appendix p 6-12).

**Figure 2.**
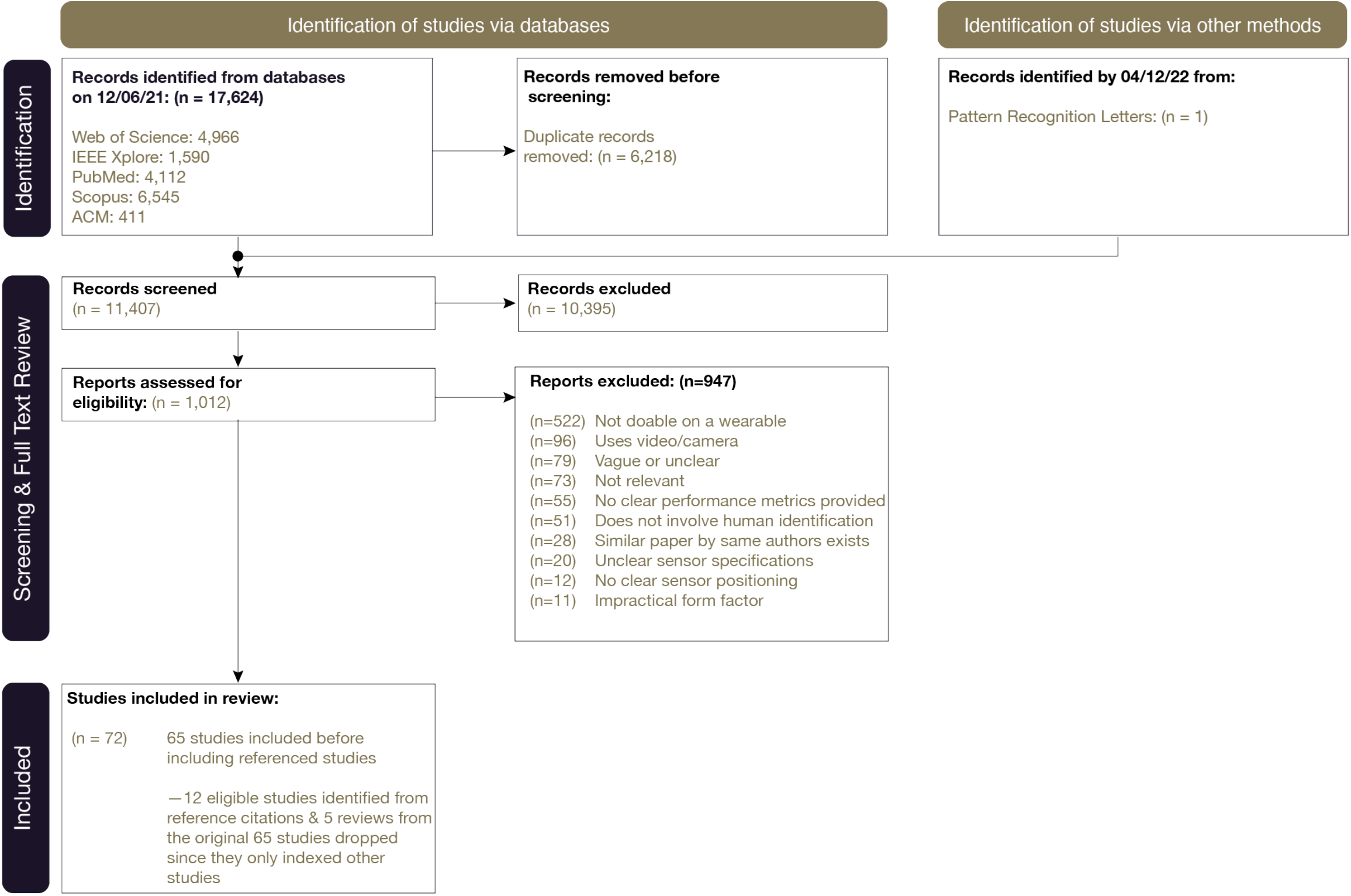
PRISMA diagram illustrating the article selection process.

For study quality assessment, standard clinical study assessment tools^21^ were not applicable because none of the included studies were clinical. Instead, we designed a custom assessment tool with four overall quality categories: high, medium, low, and very low (appendix p 13). Of the retrieved studies, sixty-seven (93%) were classified as high quality, five (7%) were medium quality, and none were low quality. We detected no publication bias^22^ in any of the included studies (appendix p 13).

Of the included studies, 20 unique sensing modalities were mentioned (Figure 3), the top three of which were electroencephalogram (EEG: N=17), inertial measurement unit (IMU: N=15), and electrocardiogram (ECG: N=8). Despite the abundance of PPG-enabled smartwatches^23^, our search revealed less investigation on PPG (N=4) as compared to ECG (N=8), and our broader search revealed the same pattern, with 297 papers on PPG and 775 on ECG in the initial search. Furthermore, in addition to studies using common sensing modalities, there were a handful of studies using less common biosignals such as seismocardiogram and bioimpedance, pointing to the importance of privacy considerations even in emerging sensing technologies.

**Figure 3.**
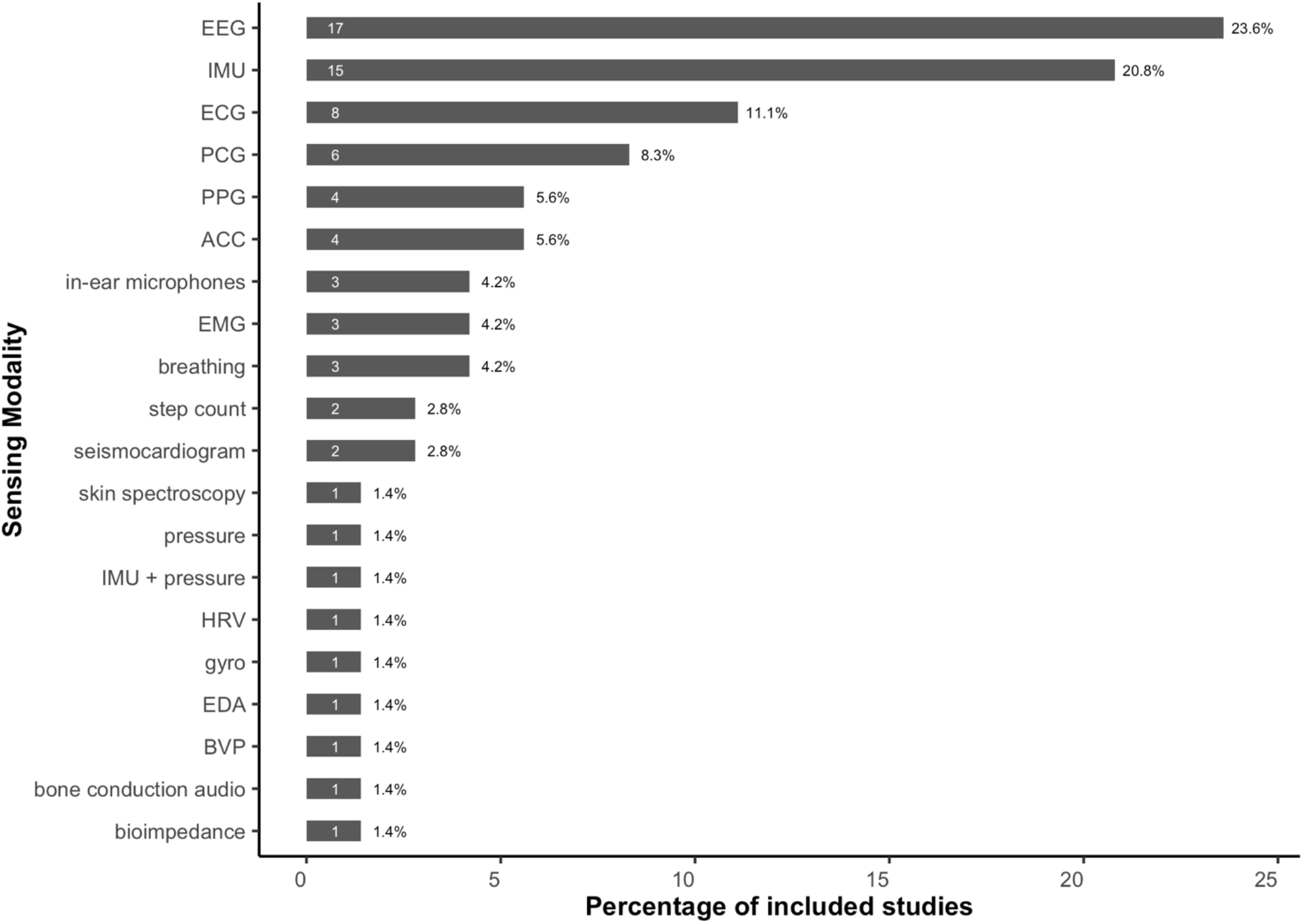
Frequency (white number inside bars) is the count of sensing modalities that are examined in the corpus of the 72 papers explored, while the percentage is the proportion of papers covering that sensing modality. Some papers explored multiple sensing modalities; hence the number of all sensing modalities (76) is more than the number of papers explored (72). Here, IMU involves the simultaneous use of an accelerometer and gyroscope, and does not include a magnetometer.

Not all 18 study characteristics were present in every paper (appendix p 1); however, every included study reported biometric identification performance (how well the system performed on the task of identifying individuals), which was this review’s key variable of interest. Because 57 (79%) of the papers used correct identification rate (CIR) as the biometric performance metric, we focus our findings on CIR; however, there are other widely-accepted biometric performance metrics, such as the equal error rate (EER), which was reported by 22 (31%) of the included studies. Notably, of the 25 studies that reported participants’ health status, all but one participant^24^ were reported to be healthy. The unhealthy participant had a heart condition which reportedly made their identification easier.^24^

We analyzed the on-body positioning of all wearables used (Figure 4). The majority of the devices were positioned on the wrist (26), head (16), or chest (13), and some of the sensing modalities were tested on multiple on-body locations (e.g., ECG was measured using sensors behind the ear, on the upper arm, on the chest, or on the wrist). In addition, two studies explored how on-body wearable device placement affects re-identification. Noh et al.^25^ found that the bioimpedance CIR was higher at the wrist (95·7%) than at the finger (77·6%), while Zhang et al.^26^ found the ECG CIR to be higher using measurements from a single arm (98·8%) as compared with using electrodes next to each ear (91·1%).

**Figure 4.**
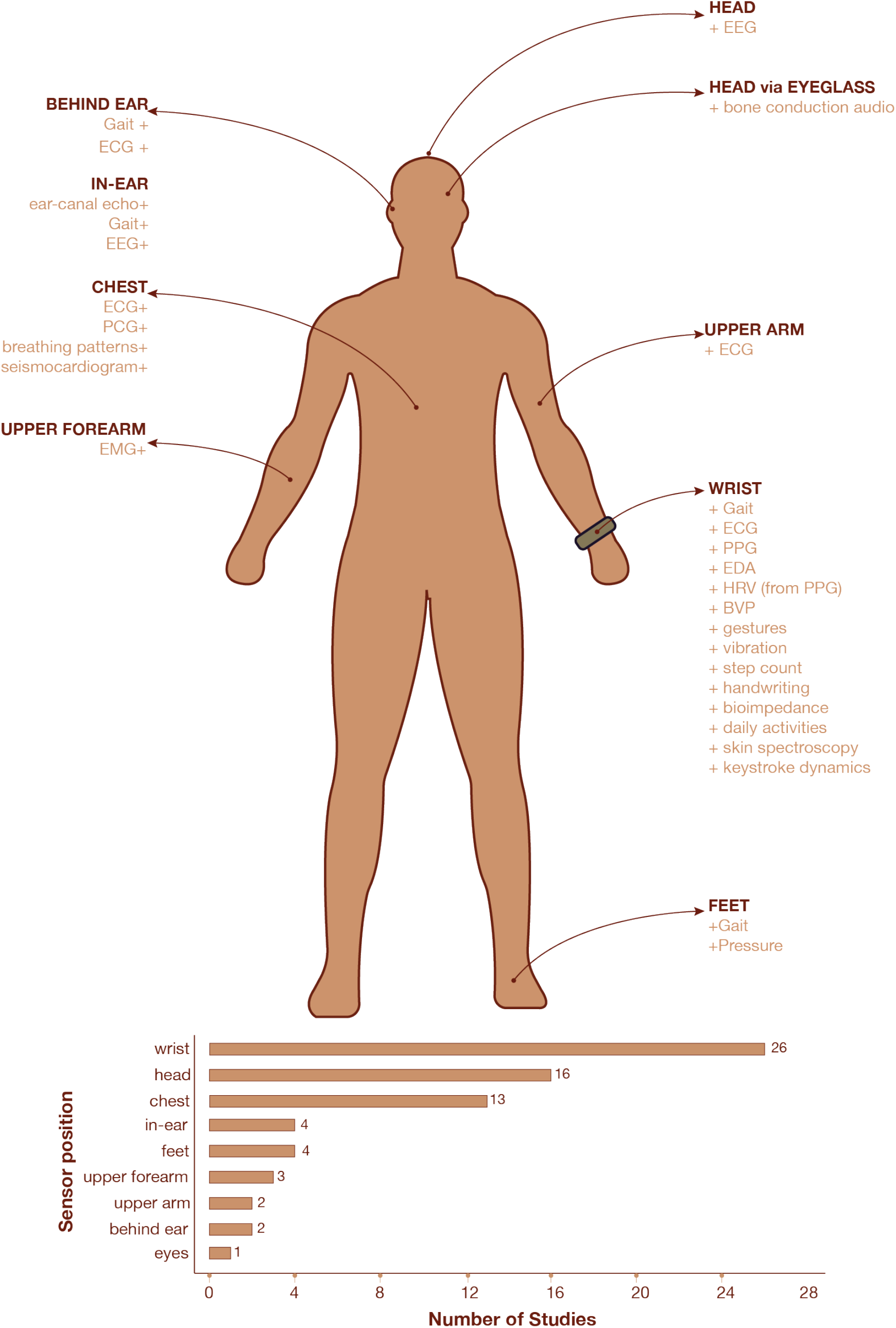
Sensor positions for included studies. This illustration excludes two studies that performed biometric identification using breathing sounds from a smartphone held in participants’ hands.

We explored the biometric identification performance of studies with the highest number of subjects for each sensing modality (appendix p 2). We also explored the minimum data needed for re-identification (Figure 5). Unfortunately, 51% of the studies did not report this characteristic; however, those that did, revealed that minimal data is required. For example, as little as 30 seconds of typing data (accelerometer and gyroscope) could achieve a CIR of 99·2% for a 34-person participant pool.^27^

**Figure 5.**
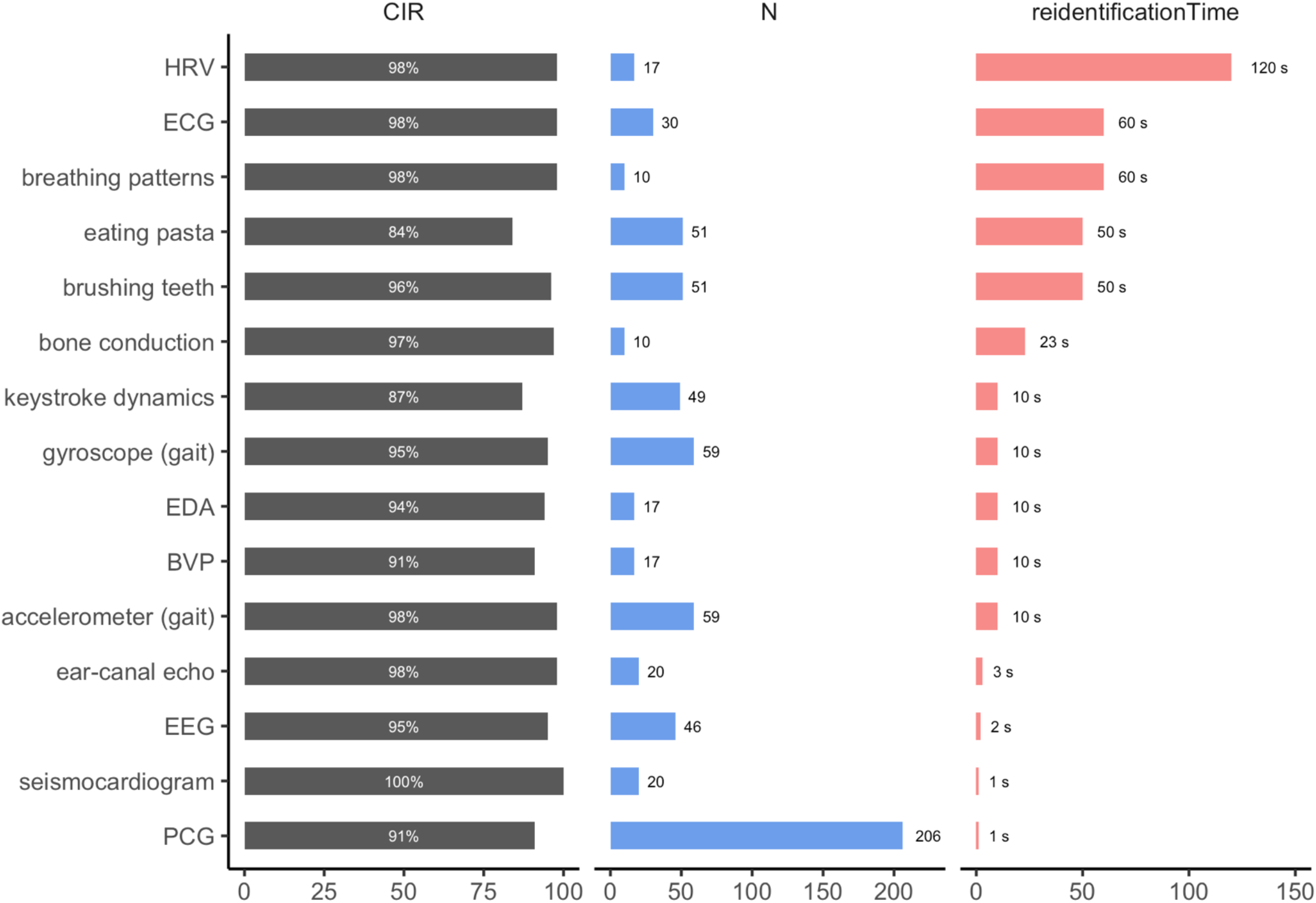
CIR, number of participants (N) alongside minimum amount of time sufficient for re-identification. The white numbers in the middle of the CIR plot represent the CIR rounded to the nearest whole number. The numbers at the end of the number of participants plot represent the number of participants. Finally, the numbers at the end of the re-identification time plot represent the minimum amount of time sufficient for re-identification (in seconds).

Given that EEG, IMU, and ECG had the largest bodies of evidence on re-identification potential, we summarized findings from these three sensor modalities. Though re-identification using IMUs has been explored when individuals perform activities of daily living such as eating^28^, brushing one’s teeth^28^, or typing^27,29^, over 50% of the studies that used IMUs focused on gait. Accordingly, we expound on gait below, and the appendix (p 10) provides tables of study characteristics of other included studies utilizing IMUs that focused on aspects of movement other than gait.

### EEG

Seventeen studies demonstrated an ability to identify an individual using EEG (average group size=20; median =16; range 4-60). Five papers (29%) reported the recording length used for re-identification, which was 21 s on average, with a median of 12·8 s. Eleven of the studies (65%) reported the health status of participants (all were healthy and aged 18-40). Activities during signal acquisition included listening to one’s favorite music, resting with eyes open or closed, cognitive loading tasks, imagined speech, and visual stimuli. The highest recorded CIR was 99·42% using the Muse 4-electrode EEG headset while participants (N=20) listened to their favorite songs.^30^ The system that could enable re-identification with the least amount of data was the Neurosky Mindwave 1-electrode EEG, which was used during resting with eyes open (N=46) and achieved a CIR of 95·48% with just two seconds of recorded data.^26^

#### ECG

Eight studies demonstrated an ability to identify an individual using only an ECG signal (average group size=15; median =10; range 5-33). Three of the studies (38%) reported the health status of participants. All participants were healthy except for a cardiopathic male aged 60 in the Randazzo et al. study, which used a custom ECG watch (1 lead, 1 kHz) to monitor 6 participants over an unspecified period during which they captured 20-63 ECGs per participant.^24^ The overall CIR of the study was 99%, and the 60-year-old cardiopathic male was reported to be the easiest to identify (CIR=100%). A separate study using the VitalJacket® (1 lead, 200 Hz) attained nearly 100% CIR for five firefighters using single heartbeats collected between 5 hours and 6 months after the training data.^31^ Even with six months between training and testing data, the proposed system could still identify all five firefighters with 100% or near-100% CIR. Finally, the most extensive study (N=33)^32^ used 1-lead OMSignal apparel over 6 weeks in free-living environments. With just 10 heartbeats, the study team’s algorithm could identify an individual with a CIR of 95·95%.

#### Gait

We define *gait* as an individual’s way of walking. Thirteen studies demonstrated an ability to identify an individual using only gait signals (average group size=34; median =30; range 8-60). However, only one of the studies reported the health status of its participants, who were all healthy. Twelve studies combined the accelerometer and gyroscope, which we refer to as an IMU, or used each sensor independently. Additionally, one study used an in-ear microphone to measure gait from walking sounds propagated through the human musculoskeletal system. One of the challenges in gait studies was the presence of multiple definitions of movement, e.g., fixed time durations, step cycles, and walk cycles, thus making it difficult to compare results across studies. However, in one study of note (N=30), just 10 seconds of data from an IMU (100 Hz accelerometer and gyroscope) from the MetaWear C Board wristband was sufficient to identify an individual with 100% CIR.^33^

## Discussion

This study reviewed a vast literature base and summarized 72 included studies. All but four of the included studies that reported CIR (N=57) demonstrated high CIR values (86-100%), suggesting that re-identification risks from wearable device data are higher than previously appreciated. Moreover, the minimum data duration for re-identification ranged from 1-300 seconds, suggesting that very small amounts of data may be sufficient to pose a privacy risk in seemingly anonymized biosensor data. All but four studies had fewer than 100 subjects; thus, it remains to be seen whether these results would scale with larger populations. The few studies with larger participant pools (N=206 to 421; 4 studies) show results consistent with those with fewer subjects (N=3 to 73; 68 studies), indicating that re-identification risks may remain a threat in larger group sizes. Further research is needed to determine to what extent large datasets pose similar risks for re-identification and what appropriate mitigation strategies are needed to protect privacy in large public biosensor databases.

This review also highlights that, in many cases, re-identification requires very little data. For example, in a study with 46 participants, 2 seconds of EEG recording could identify an individual with a CIR of 95%, and in another with 51 participants who were brushing teeth while wearing an LG G watch, 50 seconds of accelerometer and gyroscope data could identify an individual with a CIR of 96% (Figure 5). This discovery is concerning since publicly available data is becoming increasingly abundant, especially given recent data sharing advocacy and policy by influential bodies such as the US FDA^34^ and NIH.^5^ We are also strong proponents of open science and open data to enable FAIR^6^ research principles and diverse representation. Thus, we find these results to be of concern and aim, with this review, to bring the community together to explore and discuss best practices to balance the potential risks and benefits of sharing versus not sharing data (Figure 6). Consequently, as a community, we must continue reevaluating data sharing policies in the context of privacy and FAIR science principles as new research becomes available on risks and benefits on both sides.

**Figure 6.**
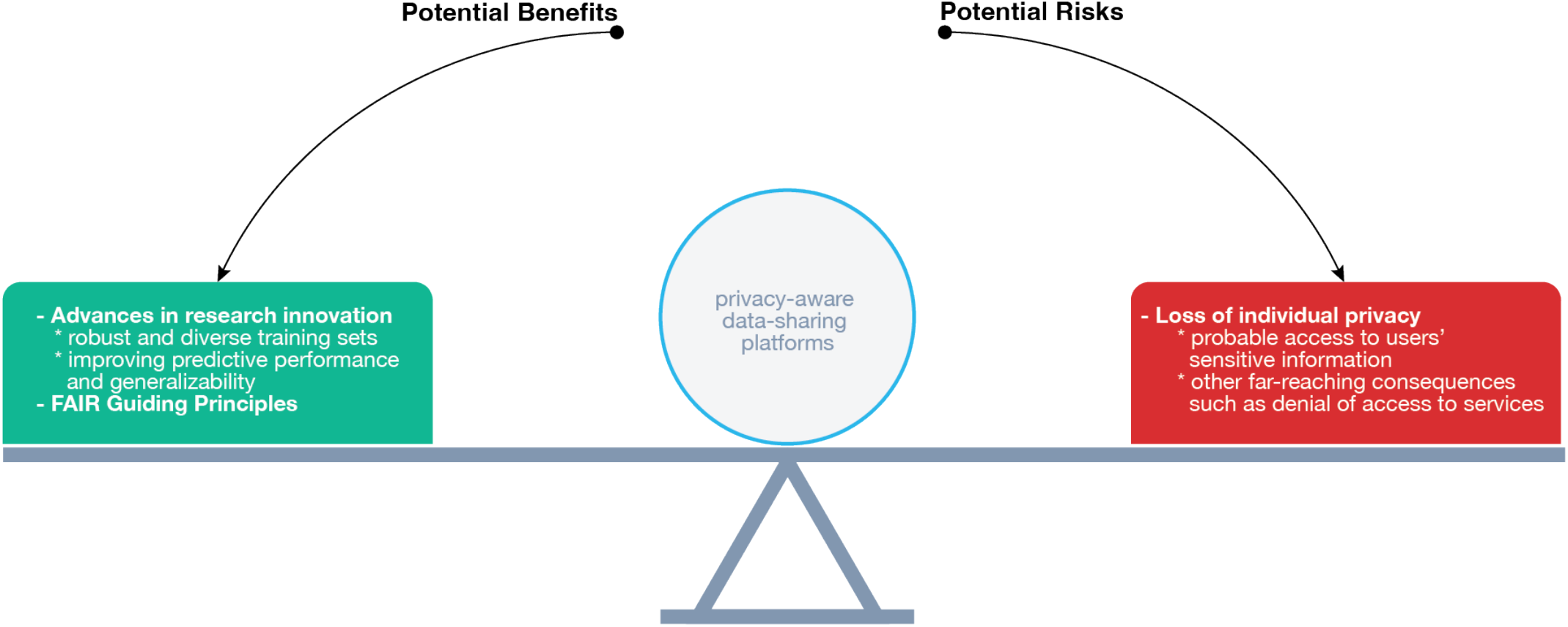
Potential benefits and potential risks of wearable data sharing can tilt the cost-benefit balance either way. Privacy-aware data sharing platforms can help to balance the risks and benefits.

In general, our findings align with similar research on state-of-the-art non-wearable devices. For example, 12-lead ECG data from two open access databases combined with other electronic health records data from 40,000 patients revealed^15^ CIR rates in similar ranges to those reported from studies using wearable ECGs. The researchers looked at 37 heart conditions, including supraventricular tachycardia, ST depression, and pacing rhythm, and recorded an overall CIR of 94·56%. The CIR for individual conditions ranged from 90·32-98·55% in all but seven conditions. Patients with premature ventricular contractions had the lowest CIR—78·54%.

In addition, 58% of the studies we analyzed utilized head-worn and wrist-worn wearables (Figure 4). This observation aligns with the global forecast for the wearable technology market^1^ for 2026, which projects head-worn and wrist-worn wearables to experience the most growth compared to other wearables.

Ultimately, there is a need to possess identifiers to re-identify someone, so merely matching individuals in de-identified/anonymized datasets does not constitute true re-identification. Re-identification concerns have been historically dismissed because the probability that an attacker gains access to data containing identifiers is low. However, an increasing number of companies are entering third-party data sharing agreements^35^, some of which are ethically tenuous^35,36^ (e.g., driven by profits, personal benefit, or political gain over public good). The desire to know more about the patient or the customer and personalize goods and services by direct advertising is a likely culprit in re-identification attempts. For example, web-scraping could reveal an individual’s medical diagnoses and personality traits, which could be used to personalize advertising or reveal more information about the subjects to benefit the re-identifying entity (Figure 1).

The findings here should not be used to justify blocking wearable biometric data sharing. On the contrary, this work exposes the need for more careful consideration of *how* data should be shared. It suggests that privacy-preserving methods will be needed for open science to flourish. For example, there is an opportunity for regulatory bodies and funding agencies to expand support for privacy-conscious data sharing platforms that mitigate re-identification risk. Such platforms could be, for instance, semi-public, research-focused data sharing platforms that only appropriately trained and approved researchers can access through two-way authentication schemes using organizational email addresses (e.g., PhysioNet^37^ and AllOfUs^38^). It should be noted that systems like this could delay or even discourage some forms of citizen science.

On a different note, none of the studies we reviewed addressed the question: in the absence of any identifying information about a group, is it possible to re-identify a person from that group using biosensor data alone? All included studies had a complete list of participants, which will not always be the case in many real-world scenarios. Therefore, there is a fundamental distinction between finding out which of the N study participants has the biometric signature of participant X versus obtaining X’s name and phone number without knowing who was in the study. In the case of genetic data, this is possible.^39^ Could future investigators merge wearable public data with public genetic data to re-identify participants? Further study is needed to determine how these concepts apply to data from wearables.

Another limitation of this review is that most studies had short session intervals or collected all data in one session. The lack of long session intervals or the collection of all data in one session prevents drawing conclusions about template aging (expected increases in error over time due to intra-individual changes, for example, changes in voice or face with age).^40,41^ Because of template aging, it might not be possible to identify individuals using widely temporally spaced data. Knowing maximum temporal intervals for any sensing modality with the ability to biometrically identify individuals could be an essential tool for policymakers. Once this is known, certain kinds of biometric data could be released to the public after scientifically determined temporal intervals. With improving algorithms, these intervals may extend as well.

The included studies had substantial missing data (appendix p 1). For example, only 35% of studies mentioned anything about participants’ health status. Of those, only one participant was unhealthy, so the results from this review might not be fully extensible to the broader population. On the other hand, if a disease is uncommon and easily identified with a biosensor, re-identifying an individual from the said sensor data would be more straightforward. Future research should explore how health status affects biometric identification. We did not evaluate multimodal re-identification techniques in this review; however, we anticipate multimodal re-identification to become more relevant in the near future.

Finally, a real risk for re-identification exists in wearable sensor data. While this risk can be minimized, it cannot be fully mitigated. Our findings reveal that basic practices of withholding identifiers from public repositories may not be sufficient to ensure privacy. More study is needed to guide the creation of policies and procedures that are sufficient to protect privacy, given the prevalence of wearable data collection and sharing. In conclusion, hope is not lost. The risk of not sharing data may be even greater than the risk of re-identification (e.g., algorithmic bias^42,43^ and failure to develop new algorithmic tools that could save lives), but new solutions are possible to reduce the risk of re-identification. For example, emphasis on research directions for methods development on privacy-protecting methods such as federated learning^44^, differential privacy^45^, and use of synthetic data^46^ could allow the community to continue to reap the many benefits of data sharing while protecting the privacy of data donors.

## Data Availability

All data produced in the present work are contained in the manuscript

## Abbreviations

N: number of subjects
EER: Equal Error Rate
CIR: Correct Identification Rate
IMU: inertial measurement unit

## Contributors

JD conceived the study. JD, DMG, LC, and YM designed the search strategy. LC retrieved all searches from the databases and exported them to Covidence. LC, YM, and DB participated in the title and abstract screening phase. LC, YM, MW, and DB participated in the full-text review and data extraction phases. LC created all tables, graphs, and other visual representations, and YM and MW participated in the quality assurance process for all tables, graphs, and visual representations. LC drafted the manuscript. All other authors read, edited, and approved the final manuscript.

## Data sharing

### All data collected in this study are available in the appendices

The search strategy is available on pages 2-5, and the list of extracted study characteristics for each included study is available on pages 5-6. We also provide a detailed list of included studies highlighting study characteristics such as sensor used, sensor positioning, and sensor resolution on pages 6-12.

### Funding

DG is funded by NIH NINDS KL2TR002542. No funding was received for this study.

## Acknowledgments

The authors would like to express their gratitude to the engineering and computer science librarian at Duke University—Sarah Park.

## References

1. Wearable Technology Market - Global Forecast to 2026 [Internet]. Markets and Markets. [cited 2022 Apr 10]. Available from: https://www.marketsandmarkets.com/Market-Reports/wearable-electronics-market-983.html?gclid=Cj0KCQjwgMqSBhDCARIsAIIVN1V0sqrk6SpYSga3rcDtWcwh8npZ08L0_s4X91gh7yPAa6QmsctB-lMaAlpqEALw_wcB

2. Cheung CC, Krahn AD, Andrade JG. The Emerging Role of Wearable Technologies in Detection of Arrhythmia. Can J Cardiol. 2018 Aug 1;34(8):1083–7.

3. Cheong SHR, Ng YJX, Lau Y, Lau ST. Wearable technology for early detection of COVID-19: A systematic scoping review. Prev Med. 2022 Sep;162:107170.

4. You L, Xu H, Zhang Q, Li T, Li C, Chen Y, et al. JDap: Supporting in-memory data persistence in javascript using Intel’s PMDK. J Syst Archit. 2019 Dec 1;101:101662.

5. NIH Data Sharing Information - Main Page [Internet]. [cited 2022 Mar 28]. Available from: https://grants.nih.gov/grants/policy/data_sharing/

6. Wilkinson MD, Dumontier M, Aalbersberg IjJ, Appleton G, Axton M, Baak A, et al. The FAIR Guiding Principles for scientific data management and stewardship. Sci Data. 2016 Mar 15;3(1):160018.

7. Seh AH, Zarour M, Alenezi M, Sarkar AK, Agrawal A, Kumar R, et al. Healthcare Data Breaches: Insights and Implications. Healthcare. 2020 May 13;8(2):133.

8. Greely HT. The Uneasy Ethical and Legal Underpinnings of Large-Scale Genomic Biobanks. Annu Rev Genomics Hum Genet. 2007 Sep 1;8(1):343–64.

9. Sweeney L. Weaving Technology and Policy Together to Maintain Confidentiality. J Law Med Ethics. 1997;25(2– 3):98–110.

10. Waldo A. A Preliminary Staff Report on “Protecting Consumer Privacy in an Era of Rapid Change: A Proposed Framework for Businesses and Policymakers” [Internet]. Available from: https://www.ftc.gov/sites/default/files/documents/public_comments/preliminary-ftc-staff-report-protecting-consumer-privacy-era-rapid-change-proposed-framework/00300-57631.pdf

11. Richens JG, Lee CM, Johri S. Improving the accuracy of medical diagnosis with causal machine learning. Nat Commun. 2020 Aug 11;11(1):3923.

12. Shatte ABR, Hutchinson DM, Teague SJ. Machine learning in mental health: a scoping review of methods and applications. Psychol Med. 2019 Jul;49(09):1426–48.

13. Mehta Y, Majumder N, Gelbukh A, Cambria E. Recent trends in deep learning based personality detection. Artif Intell Rev. 2020 Apr 1;53(4):2313–39.

14. Bota PJ, Wang C, Fred ALN, Plácido Da Silva H. A Review, Current Challenges, and Future Possibilities on Emotion Recognition Using Machine Learning and Physiological Signals. IEEE Access. 2019;7:140990–1020.

15. Ghazarian A, Zheng J, El-Askary H, Chu H, Fu G, Rakovski C. Increased Risks of Re-identification For Patients Posed by Deep Learning-Based ECG Identification Algorithms. In: 2021 43rd Annual International Conference of the IEEE Engineering in Medicine & Biology Society (EMBC). 2021. p. 1969–75.

16. Page MJ, McKenzie JE, Bossuyt PM, Boutron I, Hoffmann TC, Mulrow CD, et al. The PRISMA 2020 statement: an updated guideline for reporting systematic reviews. BMJ. 2021 Mar 29;n71.

17. Maiorana E. A survey on biometric recognition using wearable devices. Pattern Recognit Lett. 2022 Apr 1;156:29–37.

18. Covidence - Better systematic review management [Internet]. Covidence. [cited 2022 Jun 1]. Available from: https://www.covidence.org/

19. Costache A, Badescu E, Popescu D, Ichim L. Identifying Persons from Iris Image. In: 2021 13th International Conference on Electronics, Computers and Artificial Intelligence (ECAI). 2021. p. 1–4.

20. Qin Z, Huang G, Xiong H, Qin Z, Choo KKR. A Fuzzy Authentication System Based on Neural Network Learning and Extreme Value Statistics. IEEE Trans Fuzzy Syst. 2021 Mar;29(3):549–59.

21. Brożek JL, Akl EA, Alonso-Coello P, Lang D, Jaeschke R, Williams JW, et al. Grading quality of evidence and strength of recommendations in clinical practice guidelines. Allergy. 2009 May;64(5):669–77.

22. Guyatt GH, Oxman AD, Montori V, Vist G, Kunz R, Brozek J, et al. GRADE guidelines: 5. Rating the quality of evidence—publication bias. J Clin Epidemiol. 2011 Dec;64(12):1277–82.

23. Kwon S, Kim H, Yeo WH. Recent advances in wearable sensors and portable electronics for sleep monitoring. iScience. 2021 May 21;24(5):102461.

24. Randazzo V, Cirrincione G, Pasero E. Shallow Neural Network for Biometrics from the ECG-WATCH. In: Intelligent Computing Theories and Application: 16th International Conference, ICIC 2020, Bari, Italy, October 2–5, 2020, Proceedings, Part I [Internet]. Berlin, Heidelberg: Springer-Verlag; 2020 [cited 2022 May 19]. p. 259–69. Available from: https://doi.org/10.1007/978-3-030-60799-9_22

25. Noh HW, Sim JY, Ahn CG, Ku Y. Electrical Impedance of Upper Limb Enables Robust Wearable Identity Recognition against Variation in Finger Placement and Environmental Factors. Biosensors. 2021 Oct;11(10):398.

26. Zhang R, Yan B, Tong L, Shu J, Song X, Zeng Y. Identity Authentication Using Portable Electroencephalography Signals in Resting States. IEEE Access. 2019;7:160671–82.

27. Acar A, Aksu H, Uluagac AS, Akkaya K. A Usable and Robust Continuous Authentication Framework Using Wearables. IEEE Trans Mob Comput. 2021 Jun;20(6):2140–53.

28. Weiss GM, Yoneda K, Hayajneh T. Smartphone and Smartwatch-Based Biometrics Using Activities of Daily Living. IEEE Access. 2019;7:133190–202.

29. Rahman KA, Alam N, Musarrat J, Madarapu A, Hossain MS. Smartwatch Dynamics: A Novel Modality and Solution to Attacks on Cyber-behavioral Biometrics for Continuous Verification? In: 2020 International Symposium on Networks, Computers and Communications (ISNCC). 2020. p. 1–5.

30. Sooriyaarachchi J, Seneviratne S, Thilakarathna K, Zomaya AY. MusicID: A Brainwave-based User Authentication System for Internet of Things [Internet]. arXiv; 2020 [cited 2022 May 19]. Available from: http://arxiv.org/abs/2006.01751

31. Ye C, Kumar BVKV, Coimbra MT. Human identification based on ECG signals from wearable health monitoring devices. In: Proceedings of the 4th International Symposium on Applied Sciences in Biomedical and Communication Technologies - ISABEL ‘11 [Internet]. Barcelona, Spain: ACM Press; 2011 [cited 2022 May 19]. p. 1–5. Available from: http://dl.acm.org/citation.cfm?doid=2093698.2093723

32. Bahareh Pourbabaee, Howe-Patterson M, Reiher E, Benard F. Deep Convolutional Neural Network for ECG-based Human Identification. CMBES Proc [Internet]. 2018 May 8 [cited 2022 May 20];41. Available from: https://proceedings.cmbes.ca/index.php/proceedings/article/view/684

33. Sudhakar SRV, Kayastha N, Sha K. ActID: An efficient framework for activity sensor based user identification. Comput Secur. 2021 Sep 1;108:102319.

34. Platt R, Brown JS, Robb M, McClellan M, Ball R, Nguyen MD, et al. The FDA Sentinel Initiative — An Evolving National Resource. N Engl J Med. 2018 Nov 29;379(22):2091–3.

35. Fuller M. Big data and the Facebook scandal: Issues and responses. Theology. 2019 Jan;122(1):14–21.

36. Schneble CO, Elger BS, Shaw D. The Cambridge Analytica affair and Internet-mediated research. EMBO Rep [Internet]. 2018 Aug [cited 2022 Sep 24];19(8). Available from: https://onlinelibrary.wiley.com/doi/10.15252/embr.201846579

37. PhysioNet [Internet]. [cited 2022 Aug 2]. Available from: https://physionet.org/

38. All of Us Research Program | National Institutes of Health (NIH) [Internet]. All of Us Research Program | NIH. 2020 [cited 2022 Aug 2]. Available from: https://allofus.nih.gov/future-health-begins-all-us

39. Identifying Personal Genomes by Surname Inference [Internet]. [cited 2022 Jun 19]. Available from: https://www.science.org/doi/10.1126/science.1229566

40. Manjani I, Sumerkan H, Flynn PJ, Bowyer KW. Template aging in 3D and 2D face recognition. In: 2016 IEEE 8th International Conference on Biometrics Theory, Applications and Systems (BTAS). 2016. p. 1–6.

41. Matveev Y. The Problem of Voice Template Aging in Speaker Recognition Systems. In: Železný M, Habernal I, Ronzhin A, editors. Speech and Computer [Internet]. Cham: Springer International Publishing; 2013 [cited 2022 Jun 19]. p. 345–53. (Hutchison D, Kanade T, Kittler J, Kleinberg JM, Mattern F, Mitchell JC, et al., editors. Lecture Notes in Computer Science; vol. 8113). Available from: http://link.springer.com/10.1007/978-3-319-01931-4_46

42. Starke G, De Clercq E, Elger BS. Towards a pragmatist dealing with algorithmic bias in medical machine learning. Med Health Care Philos. 2021 Sep;24(3):341–9.

43. Vayena E, Blasimme A, Cohen IG. Machine learning in medicine: Addressing ethical challenges. PLOS Med. 2018 Nov 6;15(11):e1002689.

44. Rieke N, Hancox J, Li W, Milletarì F, Roth HR, Albarqouni S, et al. The future of digital health with federated learning. Npj Digit Med. 2020 Dec;3(1):119.

45. Lv Z, Piccialli F. The Security of Medical Data on Internet Based on Differential Privacy Technology. ACM Trans Internet Technol. 2021 Jun 15;21(3):1–18.

46. Alzantot M, Chakraborty S, Srivastava M. SenseGen: A deep learning architecture for synthetic sensor data generation. In: 2017 IEEE International Conference on Pervasive Computing and Communications Workshops (PerCom Workshops). 2017. p. 188–93.

