## Supplementary Materials for "Does de-identification of data from wearable Biometric Monitoring Technologies give us a false sense of security? A systematic review"

#### Appendix I: Figures

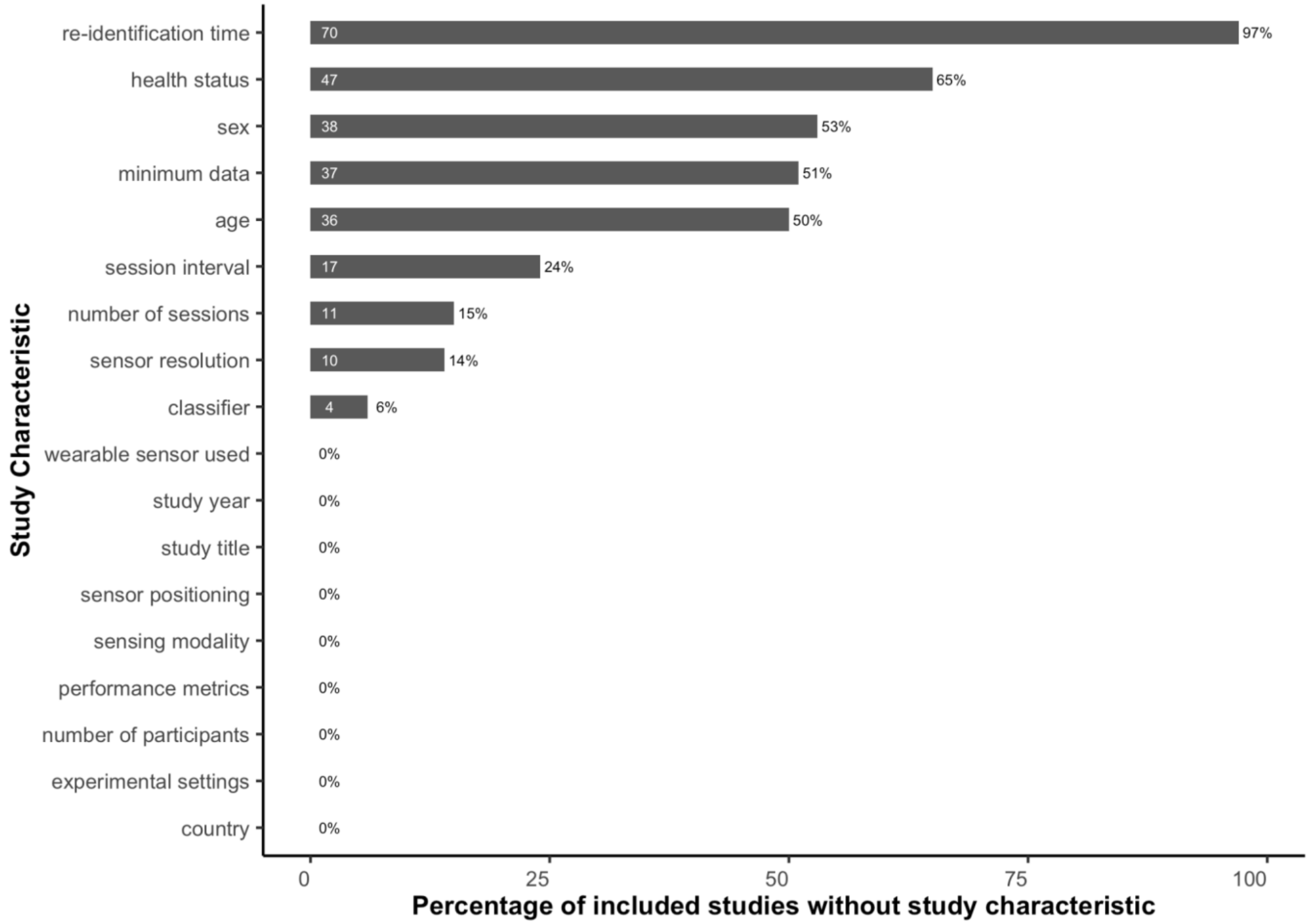


**Supplementary Figure 1.** Study characteristics missingness

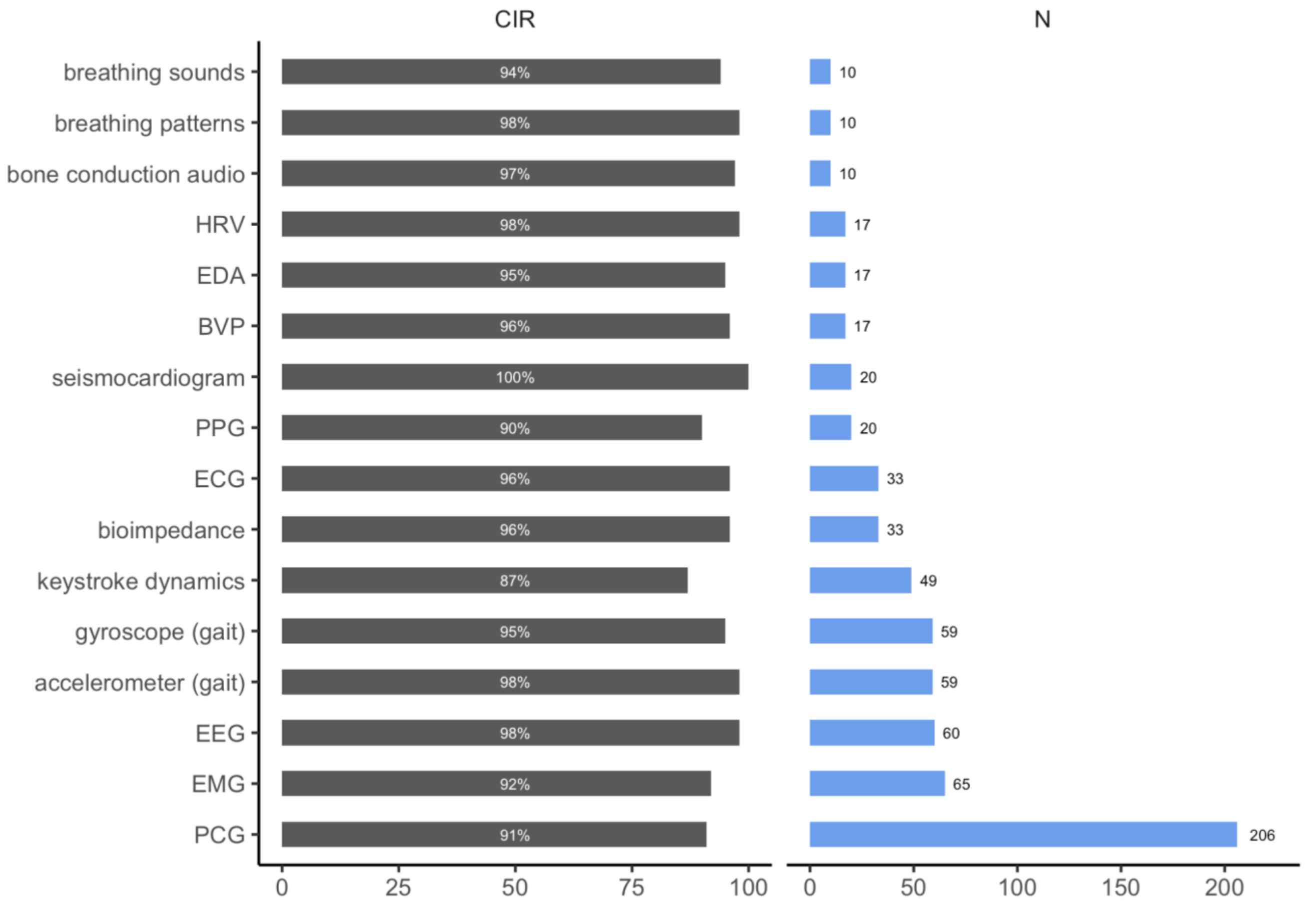


**Supplementary Figure 2.** CIR alongside the highest number of subjects (N)**.** The white numbers in the middle of the CIR plot represent the CIR rounded to the nearest whole number. The numbers at the end of the number of participants plot represent the highest number of participants for that sensing modality.

#### Appendix II: Search Strategy

##### Web of Science

Topic = ((re-id* OR reid* OR identi*) AND

(biometric* OR biosensor*) AND

(acceleromet* OR gyroscope OR magnetomet* OR IMU OR “inertial measurement unit” OR gait OR altimet* OR “pulse transit time” OR PTT OR “blood pressure” OR SpO2 OR oximet* OR photoplethysmogra* OR PPG OR O2 OR oxygen OR respiromet* OR VO2* OR “heart rate” OR “heartrate” OR cardiogra* OR electrocardiogra* OR ECG OR EKG OR seismocardiogra* OR ballistocardiogra* OR phonocardiogra* OR electromyogra* OR EMG OR pulse OR “bio-radar*” OR bioradar* OR Doppler OR respiration OR “lung sound” OR “breath sound” OR acoustic* OR electroencephalogra* OR EEG OR “galvanic skin response” OR GSR OR “electrodermal activity” OR EDA OR “eye movement” OR “pupil diameter” OR “pupil size” OR gesture* OR touch OR microphone OR voice OR temperature OR glucose OR “carbon dioxide” OR CO2 OR bioimpedance OR impedance OR bluetooth OR proximity OR “ambient light” OR UV OR “ultra-violet” OR “battery usage” OR battery))

**Query Link**
Link

##### Scopus

( TITLE-ABS-KEY ( re-id* OR reid* OR identi* ) AND TITLE-ABS-KEY ( biometric* OR biosensor* ) AND TITLE-ABS-KEY ( acceleromet* OR gyroscope OR magnetomet* OR imu OR "inertial measurement unit" OR gait OR altimet* OR "pulse transit time" OR ptt OR "blood pressure" OR spo2 OR oximet* OR photoplethysmogra* OR ppg OR o2 OR oxygen OR respiromet* OR vo2* OR "heart rate" OR "heartrate" OR cardiogra* OR electrocardiogra* OR ecg OR ekg OR seismocardiogra* OR ballistocardiogra* OR phonocardiogra* OR electromyogra* OR emg OR pulse OR "bio-radar*" OR bioradar* OR doppler OR respiration OR "lung sound" OR "breath sound" OR acoustic* OR electroencephalogra* OR eeg OR "galvanic skin response" OR gsr OR "electrodermal activity" OR eda OR "eye movement" OR "pupil diameter" OR "pupil size" OR gesture* OR touch OR microphone OR voice OR temperature OR glucose OR "carbon dioxide" OR co2 OR bioimpedance OR impedance OR bluetooth OR proximity OR "ambient light" OR uv OR "ultra-violet" OR "battery usage" OR battery ) )

**Query Link**
Link

##### PubMed

(biometric identification[MeSH Terms] OR ((biometric* OR biosensor*) AND (re-id* OR reid* OR identi*))) AND (acceleromet* OR gyroscope OR magnetomet* OR IMU OR “inertial measurement unit” OR gait OR altimet* OR “pulse transit time” OR PTT OR “blood pressure” OR SpO2 OR oximet* OR photoplethysmogra* OR PPG OR O2 OR oxygen OR respiromet* OR VO2 OR VO2max OR “heart rate” OR “heartrate” OR cardiogra* OR electrocardiogra* OR ECG OR EKG OR seismocardiogra* OR ballistocardiogra* OR phonocardiogra* OR electromyogra* OR EMG OR pulse OR “bio-radar*” OR bioradar* OR Doppler OR respiration OR “lung sound” OR “breath sound” OR acoustic* OR electroencephalogra* OR EEG OR “galvanic skin response” OR GSR OR “electrodermal activity” OR EDA OR “eye movement” OR “pupil diameter” OR “pupil size” OR gesture* OR touch OR microphone OR voice OR temperature OR glucose OR “carbon dioxide” OR CO2 OR bioimpedance OR impedance OR bluetooth OR proximity OR “ambient light” OR UV OR “ultra-violet” OR “battery usage” OR battery)

**Query Link**
Link

##### IEEE Xplore (search abstract)

(re-id OR reid* OR re-identi* OR reidenti* OR identi*) AND
(biometric* OR biosensor*) AND
(acceleromet* OR gyroscope OR magnetomet* OR IMU OR “inertial measurement unit” OR gait OR altimet* OR “pulse transit time” OR PTT OR “blood pressure” OR SpO2)

**Query Link (369 hits)**
Link

(re-id OR reid* OR re-identi* OR reidenti* OR identi*) AND
(biometric* OR biosensor*) AND
(oximet* OR photoplethysmogra* OR PPG OR O2 OR oxygen OR respiromet* OR VO2 OR VO2max OR “heart rate” OR “heartrate”)

**Query Link (80 hits)**
Link

(re-id OR reid* OR re-identi* OR reidenti* OR identi*) AND
(biometric* OR biosensor*) AND
(cardiogra* OR electrocardiogra* OR ECG OR EKG OR seismocardiogra*)

**Query Link (247 hits)**
Link

(re-id OR reid* OR re-identi* OR reidenti* OR identi*) AND
(biometric* OR biosensor*) AND
(ballistocardiogra* OR phonocardiogra* OR electromyogra* OR EMG OR pulse)

**Query Link (58 hits)**
Link

(re-id OR reid* OR re-identi* OR reidenti* OR identi*) AND
(biometric* OR biosensor*) AND
(“bio-radar*” OR bioradar* OR Doppler OR respiration OR “lung sound” OR “breath sound” OR acoustic* OR electroencephalogra* OR EEG OR “galvanic skin response” OR GSR OR “electrodermal activity” OR EDA OR “eye movement” OR “pupil diameter” OR “pupil size”)

**Query Link (268 hits)**
Link

(re-id OR reid* OR re-identi* OR reidenti* OR identi*) AND
(biometric* OR biosensor*) AND
(gesture* OR touch OR microphone OR voice OR temperature OR glucose OR “carbon dioxide” OR CO2 OR bioimpedance OR impedance OR bluetooth OR proximity OR “ambient light” OR UV OR “ultra-violet” OR “battery usage” OR battery)

**Query Link (568 hits)**
Link

##### ACM (search The ACM Full-Text collection)

[[Abstract: re-id*] OR [Abstract: reid*] OR [Abstract: identi*]] AND [[Abstract: biometric*] OR [Abstract: biosensor*]] AND [[All: acceleromet*] OR [All: gyroscope] OR [All: magnetomet*] OR [All: imu] OR [All: "inertial measurement unit"] OR [All: gait] OR [All: altimet*] OR [All: "pulse transit time"] OR [All: ptt] OR [All: "blood pressure"] OR [All: spo2] OR [All: oximet*] OR [All: photoplethysmogra*] OR [All: ppg] OR [All: o2] OR [All: oxygen] OR [All: respiromet*] OR [All: vo2*] OR [All: "heart rate"] OR [All: "heartrate"] OR [All: cardiogra*] OR [All: electrocardiogra*] OR [All: ecg] OR [All: ekg] OR [All: seismocardiogra*] OR [All: ballistocardiogra*] OR [All: phonocardiogra*] OR [All: electromyogra*] OR [All: emg] OR [All: pulse] OR [All: "bio-radar*"] OR [All: bioradar*] OR [All: doppler] OR [All: respiration] OR [All: "lung sound"] OR [All: "breath sound"] OR [All: acoustic*] OR [All: electroencephalogra*] OR [All: eeg] OR [All: "galvanic skin response"] OR [All: gsr] OR [All: "electrodermal activity"] OR [All: eda] OR [All: "eye movement"] OR [All: "pupil diameter"] OR [All: "pupil size"] OR [All: gesture*] OR [All: touch] OR [All: microphone] OR [All: voice] OR [All: temperature] OR [All: glucose] OR [All: "carbon dioxide"] OR [All: co2] OR [All: bioimpedance] OR [All: impedance] OR [All: bluetooth] OR [All: proximity] OR [All: "ambient light"] OR [All: uv] OR [All: "ultra-violet"] OR [All: "battery usage"] OR [All: battery]]

**Query Link (411 hits)**
[Link]

#### Appendix III: Data Extraction Characteristics

| | Extracted Study Characteristic |
|---|---|
| 1 | study title |
| 2 | year the study was published |
| 3 | country in which the study was conducted |
| 4 | number of participants |
| 5 | participants’ health status |
| 6 | participants’ age |
| 7 | participants’ sex |
| 8 | sensing modality (sensor/trait) |
| 9 | wearable sensor used |
| 10 | sensor resolution |
| 11 | the number of sessions (we define a session as a unique data collection period with no resting intervals or resting intervals of less than 15-minute each. Session intervals should only exist for studies with multiple sessions.) |
| 12 | session interval (This is the time between sessions.) |

| | |
|---|---|
| 13 | sensor positioning |
| 14 | experimental settings |
| 15 | minimum amount of data needed (This is the minimum amount of data sufficient to re-identify an individual) |
| 16 | the time needed for re-identification (this is the time the algorithm takes to process the individual's data to re-identify them) |
| 17 | classifier (this is the classifier used for identifying individuals) |
| 18 | performance metrics (In biometrics systems performance is a measure of how good the system is at identifying a person. The primary metrics often include Correct Identification Rate (CIR) and Equal Error Rate (EER). EER is a point on a biometric system's Receiver Operating Characteristic (ROC) curve where the False Acceptance Rate (FAR) and the False Rejection Rate (FRR) cross. The lower the EER of a system, the better the performance.) |
| * | the stimulus used (for EEG, we extracted information on the kind of stimulus used for the study) |
| * | number of channels (for EEG, EMG and PPG we extracted information on the number of channels the device(s) used had) |
| * | number of leads (for ECG we extracted information on the number of leads for the captured ECG) |

**Table 1.** Extracted study characteristics for each included study

#### Appendix IV: Tables of Characteristics

ECG

| | Leads | N | Sessions | Session Interval | Device | Sensor Position | Resolution | Settings | Amount of data needed | Classifier | Performance |
|---|---|---|---|---|---|---|---|---|---|---|---|
| Ye et al.[1] | 1 | 5 | multiple over 6 months | - | VitalJacket® | chest | 200 Hz | free-living | 1 heartbeat | SVM | 1 heartbeat CIR: ≈62·5–98%<br>6 heartbeats CIR: ≈69–100% |
| Chandrashekhar et al.[2] | 1 | 5 | 1 | - | Apple Watch Series 5 | wrist + finger | 514 Hz | lab | - | kNN | CIR: 99·2%<br>EER: 0·8% |
| Randazzo et al.[3] | 1 | 6 | multiple | - | custom ECG Watch | between wrists | 1 kHz | lab | - | MLP | CIR: 99% |
| Zhang and Zhou[4] | 1 | 10 (arm)<br>14(ear) | 2 | - | custom single-arm ECG + ear-ECG | upper left arm + behind ear | 500 Hz | lab | - | CNN | arm CIR: 98·8%<br>ear CIR: 91·1% |
| Zhang et al.[5] | 1 | 10 | 1 | - | custom single-arm ECG | upper left arm | 500 Hz | lab | - | CNN | CIR: 98·4% |
| Lehmann and Buschek[6] | 1 | 20 | 2 (lab)<br>6 (free-living) | - | EcgMove3 & EcgMove4 | chest | 1024 Hz | lab + free-living | 3 heartbeats | RF | ROC AUC: 84·3%<br>EER: 21·91% |

| | | | | | | | | | | | |
|---|---|---|---|---|---|---|---|---|---|---|---|
| Derawi[7] | - | 30 | 1[+] | - | custom ECG | chest | 1bpm | lab | 1 minute | RBFNetwork | CIR: 97·5%<br>EER: 0·98% |
| Pourbabae et al.[8] | - | 33 | multiple over 6 weeks | - | OMsignal apparel | torso | - | free-living | 10 heartbeats | CNN | CIR: 96% |

1[+] though the paper records the sessions as 5, data was taken continuously for 5 minutes and we consider this to be 1 session

**Table 1.** Main study characteristics for included studies focusing on ECG

EEG

| | Stimulus | Channels | N | Sessions | Session Interval | Device | Sensor Position | Resolution | Settings | Amount of data needed | Classifier | Performance |
|---|---|---|---|---|---|---|---|---|---|---|---|---|
| Ericsen et al.[9] | visual | 14 | 4 | 1 | - | Emotiv EPOC | head | 128 Hz | lab | - | - | CIR: 87·5%<br>FAR: 12·5%<br>FRR: 12·5% |
| Thomas et al.[10] | REO + REC | 14 | 6 | multiple | - | Emotiv EPOC | head | 128 Hz | lab | 20 s | - | CIR: 88·33% |
| Yap et al.[11] | REC, visual | 14 | 8 | 2 (morning + afternoon sessions) | - | Emotiv EPOC+ | head | 256 Hz | lab | - | SVM | **REC** CIR: 96·42%<br>**visual** CIR: 99·06% |
| Bashar et al.[12] | REC | 5 | 9 | multiple over 6 weeks | - | Emotiv Insight | head | 128 Hz | lab | - | SVM | CIR: 94·44% |
| Jayarathne et al.[13] | REC, visual, mental task | 14 | 12 | 1 | - | Emotiv EPOC | head | 128 Hz | lab | - | kNN, QDA, SVM | CIR: 100% |
| Dan et al.[14] | REC | 1 | 13 | 3 | at least 1 day | Neurosky MindSet | head | 512 Hz | lab | - | SVM | CIR: 87·2% |
| Rosli et al.[15] | visual | 14 | 13 | 1 | - | Emotiv EPOC | head | 128 Hz | lab | - | kNN | CIR: 92·8% |
| Nakamura et al.[16] | REC | 2 | 15 | 2 | 5-15 days | custom in-ear EEG | in-ear | 1200 Hz | lab | 60 s | cosine distance | CIR: 95·7% |
| Dai et al.[17] | mental task | 1 | 16 | 1 | - | Neurosky Mindwave | head | 512 Hz | lab | 10 s | SVM | CIR: 93·73% |
| Moctezuma and Molinas[18] | imagined speech | 14 | 20 | 1 | - | Emotiv EPOC | head | 128 Hz | lab | - | SVM | CIR: 92% |
| Sooriyaarachchi et al.[19] | music with eyes closed | 4 | 20 | multiple over 2 months | > 1 week | Muse Headset | head | 220 Hz | lab | - | RF | **listening to favorite song** CIR: 99·46%<br>**listening to same song** CIR: 98·39% |
| Koike-Akino et al.[20] | mental task | 14 | 25 | 1-2 | 15-60 min | Emotiv Epoc | head | 128 Hz | naturalistic | 12.8 s | LDA | CIR: 96·7% |

| | | | | | | | | | | | | |
|---|---|---|---|---|---|---|---|---|---|---|---|---|
| Abo-Zahhad et al.[21] | eye blinks | 1 | 25 | 1-2 | > 2 weeks | Neurosky Mindwave | head | 512 Hz | lab | - | LDA | CIR: 97·3 %<br>EER: 3·7% |
| Gopal and Shukla[22] | visual + free-text typing | 14 | 26 | 2 | 4 - 5 days | Emotiv EPOC | head | 128 Hz | lab | - | deep learning | EER: 0·14% |
| Moctezuma et al.[23] | imagined speech | 14 | 27 | 1 | - | Emotiv EPOC | head | 128 Hz | lab | - | RF | CIR: 98% |
| Zhang et al.[24] | REO | 1 | 46 | 1 | - | Neurosky Mindwave | head | 512 Hz | lab | 2 s | Ensemble Classifier | CIR: 95·48% |
| Kaur et al.[25] | music with eyes closed | 14 | 60 | 1 | - | Emotiv EPOC | head | 128 Hz | lab | - | HMM | CIR: 97·5% |

**Table 2.** Main study characteristics for included studies focusing on EEG

EMG

| | Channels | N | Sessions | Session Interval | Device | Sensor Position | Resolution | Settings | Amount of data needed | Classifier | Performance |
|---|---|---|---|---|---|---|---|---|---|---|---|
| Lu et al.[26] | 8 | 21 | 1 | - | MYO armband | upper forearm | 200 Hz | lab | - | CNN | CIR: 99·20% |
| Lu et al.[27] | 8 | 21 | 1 | - | MYO armband | upper forearm | 200 Hz | lab | - | ExtraTrees Classifier | CIR: 99·21% |
| Raurale et al.[28] | 8 | 65 | multiple | - | MYO armband | upper forearm | 200 Hz | lab | - | deep learning | CIR: 92·08%<br>EER: 16·42% |

**Table 3.** Main study characteristics for included studies focusing on EMG

PPG

| | N | Sessions | Session Interval | Device | Sensor Position | Resolution | Settings | Amount of data needed | Classifier | Performance |
|---|---|---|---|---|---|---|---|---|---|---|
| Cao et al.[29] | 7 | 6 | 10 days | custom PPG wristband | wrist | 400 Hz | lab | 4 heartbeats | RF | F1 score: 97·2% |
| Jindal et al.[30] | 11 | 1 | - | wrist-worn device | wrist | 125 Hz | lab | - | RBM + DBN | CIR: 96·1% |
| Everson et al.[31] | 12 | 1 | - | wrist-worn device | wrist | 125 Hz | lab | - | CNN+LSTM | CIR: 96%<br>Precision: 89%<br>Recall: 84% |
| Zhao et al.[32] | 20 | 1 | - | custom wearable | wrist | 300 Hz | lab | - | Gradient Boosting Tree | CIR: >90% |

**Table 4.** Main study characteristics for included studies focusing on PPG

PCG

| | N | Sessions | Session Interval | Device | Sensor Position | Resolution | Settings | Amount of data needed | Classifier | Performance |
|---|---|---|---|---|---|---|---|---|---|---|
| Khan et al.[33] | 30 | multiple in 4 months | - | BSL stethoscope SS30L* | chest near pulmonary valve | - | lab | <=10 s | SVM | CIR: 95·4% |
| Zhao and Shen[34] | 30 | 3 | - | HL200 electronic stethoscope* | chest near pulmonary valve | 2000 Hz | lab | 8 s | GMM | CIR: 100% |
| Cheng et al.[35] | 40 | 2 | ≈1 hr | Ω shoulder-belt wireless heart sound sensor | chest supported by shoulder belt | 11025 Hz | lab | 1 heartbeat | Euclidean Distance and the close principle | CIR: 97·5% |
| Fahad et al.[36] | 50 | 2 | <24 hrs | ThinkLabs Rhythm digital electronic stethoscope* | chest near pulmonary valve | 11025 Hz | lab | - | Ensemble of Bagged Decision Trees | CIR: 86·7% |
| F. Beritelli[37] | 70 | 2 | 2-35 days | ThinkLabs Rhythm DS32A electronic stethoscope* | chest near pulmonary valve | 11025 Hz | lab | 6 s | linear separability curves | EER: 9% |
| Abo-Zahhad et al.[38] | 206 | 2 | <24 hrs | ThinkLabs Rhythm digital electronic stethoscope* | chest near pulmonary valve | 11025 Hz | lab | 1 s | LDA and Bayes Decision Rule | CIR: 91·05%<br>EER: 3·2% |

**Table 5.** Main study characteristics for included studies focusing on PCG

Gait

| | Sensor(s) | N | Sessions | Session Interval | Device | Sensor Position | Resolution | Settings | Amount of data needed | Classifier | Performance |
|---|---|---|---|---|---|---|---|---|---|---|---|
| Kim et al.[39] | acc + gyro | 8 | 1 | - | inertial sensor (EBIMU24gv3) | shoe | 100 Hz | lab | - | RF | CIR: 97·9%<br>EER: 2·4% |
| Jeon et al.[40] | acc + gyro | 8 | 1 | - | prototype IMU earring | behind ear | 20 Hz | lab | 5 step cycles | CNN (Alexnet) | CIR: 100% |
| Cola et al.[41] | acceleration | 15 | 1 | - | Shimmer 3 | wrist | 50 Hz | lab | - | - | AUC: 99·6%<br>EER: 2·9% |
| Retsinas et al.[42] | acc + gyro | 20 | 20+ days | 2 hr charging | Samsung Gear S3 Frontier smartwatch | wrist | 20 Hz | free-living | - | DNN | **We Thank** |
| Thank I /Tao et al.[43] | acc + gyro | 22 | 1 | - | custom inertial sensor | in-shoe | 20 Hz | controlled free-living | - | PNN | CIR 87·5% |
| Moon et al.[44] | acc + gyro + pressure | 30 | 1 | - | Footlogger insole | shoe insole | 100 Hz | lab | - | CNN + RNN | **acc** CIR: 97·98%<br>**gyro** CIR: 98·24%<br>**pressure** CIR: 98·85% |
| Sudhakar et al.[45] | acc + gyro | 30 | 2 | - | MetaWear C Board | wrist | 100 Hz | lab | 10 s | SWV-SVM | CIR: 100% |
| Ferlini et al.[46] | in-ear microphones | 31 | 8 | - | MINISO Marvel earphones | in-ear | 48 kHz | lab | - | SVM | BAC: 97·26%<br>FAR: 3·23%<br>FRR: 2·25% |

| | | | | | | | | | | | |
|---|---|---|---|---|---|---|---|---|---|---|---|
| Musale et al.[47] | acc + gyro | 51 | 1 | - | Motorola 360 Sport 2nd Gen smartwatch | wrist | 100 Hz | lab | - | RF | CIR: 91·8%<br>EER: 8·2% |
| Baek et al.[48] | acc + gyro | 51 | 1 | - | Motorola 360 Sport 2nd Gen smartwatch | wrist | 100 Hz | lab | 1 walk cycle | DNN | EER: 1·8% |
| Johnston and Weiss[49] | accelerometer | 59 | 1 | - | LG G Watch | wrist | 20 Hz | lab | 10 s | MLP | CIR: 98%<br>EER: 2% |
| Johnston and Weiss[49] | gyroscope | 59 | 1 | - | LG G Watch | wrist | 20 Hz | lab | 10 s | MLP | CIR: 94·6%<br>EER: 6·3% |
| Al-Naffakh et al.[50] | acc + gyro | 60 | multiple | 3 weeks | Microsoft Band | wrist | 32 Hz | lab | - | MLP | EER: 0·05% |

**Table 6.** Main study characteristics for included studies focusing on gait

IMU Sensors

| | Trait | N | Sessions | Session Interval | Device | Sensor Position | Resolution | Settings | Amount of data needed | Classifier | Performance |
|---|---|---|---|---|---|---|---|---|---|---|---|
| Acar et al.[51] | keystroke dynamics (acc + gyro) | 34 | 2 | - | Android Wear smartwatch | wrist | 100 Hz | lab | 30 s | MLP | CIR: 99·2%<br>EER: 1% |
| Rahman et al.[52] | keystroke dynamics (acc + gyro) | 49 | 1 | - | LG G Watch | wrist | 20 Hz | lab | 10 s | MLP | CIR: 87·2%<br>FAR: 0·2% |
| Griswold-Steiner et al.[53] | handwriting (acc) | 53 | 2 | >24 hrs | LG Urbane 2 smartwatch | wrist | 100 Hz | lab | - | CNN + RNN | EER: 5·51% |
| Buriro et al.[54] | gestures (acc + gyro) | 11 | 3 | 1 day | Motorola Moto 360 smartwatch | wrist | 50 Hz | lab | - | MLP | CIR: 80·52%<br>FAR: 21·65% |
| Weiss et al.[55] | brushing teeth (acc + gyro) | 51 | 1 | - | LG G Watch | wrist | 20 Hz | lab | 50 s | RF | CIR: 96·1%<br>EER: 14·4% |
| Weiss et al.[55] | eating pasta (acc + gyro) | 51 | 1 | - | LG G Watch | wrist | 20 Hz | lab | 50 s | RF | CIR: 84·0%<br>EER: 18·5% |
| Retsinas et al.[42] | sleeping (acc + gyro + hrm) | 20 | 20+ days | 2 hr charging | Samsung Gear S3 Frontier smartwatch | wrist | 20 Hz | free-living | - | DNN | **acc** CIR: 57·67%<br>**gyro** CIR: 67·97%<br>**hrm** CIR: 71·09% |
| Saleheen et al.[56] | accelerometer | 353 | 70 days at least 8 hrs/day | - | unspecified wrist-worn wearables | wrist | 25 Hz | free-living | - | CNN | Re-ID-Risk: 96% |
| Lee et al.[57] | vibration (acc + gyro) | 20 | 2 | 7 days | Apple Watch Series 3 | wrist | 100 Hz | lab | - | SVM | EER: 1·37% |

**Table 7.** Main study characteristics for included studies focusing on IMUs

Other Sensors

| | Trait | N | Sessions | Session Interval | Device | Sensor Position | Resolution | Settings | Amount of data needed | Classifier | Performance |
|---|---|---|---|---|---|---|---|---|---|---|---|
| Jo et al.[58] | skin spectroscopy | 73 | 1 | - | Multispectral Skin Photomatrix Device | wrist | - | lab | - | LDA | FAR: 0·29%<br>FRR: 3·54% |
| Maiorana & Massaroni[59] | seismocardiogram | 10 | 1 | - | 5 MEMS measurement units | chest | - | lab | 5 s | CNN | CIR: 99·9% |
| Hsu et al.[60] | seismocardiogram | 20 | 1 | - | Biopac MP36* | chest | 5 kHz | lab | 1 s | CNN (ResNet-50) | CIR: 100%<br>EER: 0% |
| Takeda et al.[61] | pressure | 10 | 1 | - | Octsens 8-channel wearable sensor | below feet | 100 Hz | lab | 8 steps | Euclidean distance | FAR: 0·02%<br>FRR: 0·83% |
| Chauhan et al.[62] | breathing sounds (microphone) | 10 | 3 in 7 days | >=3 days | iPhone 6* and Nexus 6P* | in hand | 8 kHz | lab | 1 breath | GMM | CIR: >94% |
| Chauhan et al.[63] | breathing sounds (microphone) | 10 | 3 in 7 days | >=3 days | iPhone 6* and Nexus 6P* | in hand | 8 kHz | lab | 1 breath | LSTM | F-score: 80-94% |
| Raji et al.[64] | breathing patterns | 10 | 1 | - | custom smart chestband | chest | - | lab | 1 minute | MLP | CIR: 98% |
| Gao et al.[65] | ear canal echo (in-ear microphone) | 20 | 1 | - | custom in-ear headphones with microphone | in-ear | 44,100 Hz | lab | 3 s | SVM | BAC: 97·52% |
| Lei et al.[66] | ear canal echo (in-ear microphone) | 50 | 1 | - | custom in-ear headphones with microphone | in-ear | - | lab | - | CNN (ResNet-18) | FAR: 5%<br>FRR: 3·2% |
| Noh et al.[67] | bioimpedance | 33 | 5 | 1 week | custom device | left wrist | - | lab | - | CNN | **upper limb** CIR: 95·70%<br>**upper limb** EER: 0·98% |
| Noh et al.[67] | bioimpedance | 33 | 5 | 1 week | custom device | left wrist | - | lab | - | CNN | **finger** CIR: 77·62%<br>**finger** EER: 5·05% |
| Schneegass et al.[68] | bone conduction audio | 10 | 1 | - | Google Glass | head via eyeglass | - | lab | 23 s | 1NN | CIR: 97%<br>EER: 6·9% |
| Piciucco et al.[69] | EDA | 17 | 2 | 7 days | Empatica E4 | wrist | 4 Hz | free-living | 10 s | CNN (MobileNet v2) | **10 s** CIR: 94·31%<br>**20 s** CIR: 94·91% |
| Piciucco et al.[69] | BVP | 17 | 2 | 7 days | Empatica E4 | wrist | 64 Hz | free-living | 10 s | CNN (MobileNet v2) | **10 s** CIR: 91·46%<br>**20 s** CIR: 95·12%<br>**30 s** CIR: 96·23% |
| Ekiz et al.[70] | HRV | 8 | 1 | - | Empatica E4 | wrist | 64 Hz | controlled free-living | 2 minutes | RF | EER: 6·77% |

| | | | | | | | | | | | |
|---|---|---|---|---|---|---|---|---|---|---|---|
| Ekiz et al.[70] | HRV | 3 | 1 | - | Samsung Gear S | wrist | 100 Hz | controlled free-living | 2 minutes | RF | EER: 13·28% |
| Ekiz et al.[70] | HRV | 17 | 1 | - | Samsung Gear S2 | wrist | 100 Hz | controlled free-living | 2 minutes | RF | CIR 98·48%<br>EER 3·96% |

**Table 8.** Main study characteristics for included studies focusing on miscellaneous sensors

Step count

| | Trait (s) | N | Sessions | Session Interval | Device | Sensor Position | Resolution | Settings | Amount of data needed | Classifier | Performance |
|---|---|---|---|---|---|---|---|---|---|---|---|
| Vhanduri and Poellabauer[71] | step counts, heart rate, calorie burn and MET | 400 | multiple over 17 months | - | Fitbit Charge HR | wrist | - | free-living | 5 minutes | SVM | **sedentary** CIR: 93%<br>**non-sedentary** CIR: 90% |
| Vhanduri and Poellabauer[72] | step counts, heart rate, calorie burn and MET | 421 | multiple over 2 years | - | Fitbit Charge HR | wrist | - | free-living | 5 minutes | SVM | CIR: 92·97% |

**Table 9.** Main study characteristics for included studies focusing on step count

#### Abbreviations

| | |
|---|---|
| N | number of subjects |
| * | study not done on a wearable |
| BAC | Balanced Accuracy |
| CIR | Correct Identification Rate |
| DBN | Deep Belief Network |
| EDA | electrodermal activity |
| FAR | False Acceptance Rate |
| GMM | Gaussian Mixture Model |
| HRV | heart rate variability |
| kNN | k-nearest neighbors |
| LSTM | Long Short-Term Memory |
| MLP | multilayer perceptron |
| REC | resting state with eyes closed |
| Re-ID Risk | Re-identification Risk |
| RNN | Recurrent Neural Network |
| SWV | Sliding Window Voting |
| RBM | Restricted Boltzmann Machines |
| K | number of sessions |
| 1NN | 1-Nearest-Neighbor |
| BVP | Blood Volume Pulse |
| CNN | Convolutional Neural Network |
| DNN | Deep Neural Network |
| EER | Equal Error Rate |
| FRR | False Rejection Rate |
| HMM | Hidden Markov Model |
| IMU | inertial measurement unit |
| LDA | Linear Discriminant Analysis |
| MET | metabolic equivalent of task |
| PNN | Probabilistic Neural Network |
| REO | resting state with eyes open |
| RF | Random Forest |
| SVM | Support Vector Machine |
| QDA | Quadratic Discriminant Analysis |

#### Appendix V: Study Quality and Publication Bias Assessment

For study quality assessment, standard clinical study assessment tools were not applicable because none of the included studies were clinical. Instead, we designed a custom assessment tool with four overall quality categories: high, medium, low, and very low.

*The following questions were multiple choice with **Yes**, **No**, or **Somewhat** as answers:*

1. Was the study objective or research question clearly stated?
2. Was the study population clearly defined, specified, and representative of the general population?
3. Was the data collected in a way that supports the research question or study objective?
4. Did the study describe the wearable device used?
5. Did the study describe the sensor resolution?
6. Did the study describe the classifier used?
7. Did the study report the amount of data needed for re-identification?
8. Were the results valid?
9. Were the results well described?

*To examine the potential of **publication bias**, we looked at each study's funding source, authors' conflicts of interest, and any other factors that could result in publication bias. We then rated each study into any of the three categories:*

- Undetected
- Strongly Suspected
- Very Strongly Suspected

*For the **overall study quality**, we classified each study into either of the four quality categories depending on the number of YES votes it received from two of the reviewers resulting in the following:*

- High Quality (7-9 YESs)
- Moderate Quality (5-6 YESs)
- Low Quality (3-4 YESs)
- Very Low Quality (0-2 YESs)

If publication bias is undetected, the overall study quality is not affected; however, when it is strongly suspected, the overall study quality degrades by one level, and when it is very strongly suspected, the overall study quality degrades by two levels.

### REFERENCES


1. Ye C, Kumar BVKV, Coimbra MT. Human identification based on ECG signals from wearable health monitoring devices. In: Proceedings of the 4th International Symposium on Applied Sciences in Biomedical and Communication Technologies - ISABEL ’11 [Internet]. Barcelona, Spain: ACM Press; 2011 [cited 2022 May 19]. p. 1–5. Available from: http://dl.acm.org/citation.cfm?doid=2093698.2093723
2. Chandrashekhar V, Singh P, Paralkar M, Tonguz OK. Pulse ID: The Case for Robustness of ECG as a Biometric Identifier. In: 2020 IEEE 30th International Workshop on Machine Learning for Signal Processing (MLSP). 2020. p. 1–6.
3. Randazzo V, Cirrincione G, Pasero E. Shallow Neural Network for Biometrics from the ECG-WATCH. In: Intelligent Computing Theories and Application: 16th International Conference, ICIC 2020, Bari, Italy, October 2–5, 2020, Proceedings, Part I [Internet]. Berlin, Heidelberg: Springer-Verlag; 2020 [cited 2022 May 19]. p. 259–69. Available from: https://doi.org/10.1007/978-3-030-60799-9_22
4. Zhang Q, Zhou D. Deep Arm/Ear-ECG Image Learning for Highly Wearable Biometric Human Identification. Ann Biomed Eng. 2018 Jan;46(1):122–34.
5. Zhang Q, Zhou D, Zeng X. PulsePrint: Single-arm-ECG biometric human identification using deep learning. In: 2017 IEEE 8th Annual Ubiquitous Computing, Electronics and Mobile Communication Conference (UEMCON). 2017. p. 452–6.
6. Lehmann F, Buschek D. Heartbeats in the Wild: A Field Study Exploring ECG Biometrics in Everyday Life. In: Proceedings of the 2020 CHI Conference on Human Factors in Computing Systems [Internet]. New York, NY, USA: Association for Computing Machinery; 2020 [cited 2022 May 19]. p. 1–14. Available from: https://doi.org/10.1145/3313831.3376536
7. Derawi M. Wireless Chest-Based ECG Biometrics. In: Park JJ (Jong H, Stojmenovic I, Jeong HY, Yi G, editors. Computer Science and its Applications. Berlin, Heidelberg: Springer; 2015. p. 567–79. (Lecture Notes in Electrical Engineering).
8. Bahareh Pourbabaee, Howe-Patterson M, Reiher E, Benard F. Deep Convolutional Neural Network for ECG-based Human Identification. CMBES Proc [Internet]. 2018 May 8 [cited 2022 May 20];41. Available from: https://proceedings.cmbes.ca/index.php/proceedings/article/view/684
9. Ericsen, Thomas KP, Vinod AP. Eeg-based biometrie authentication using self-referential visual stimuli. In: 2017 IEEE International Conference on Systems, Man, and Cybernetics (SMC). 2017. p. 3048–53.
10. Thomas KP, Vinod AP, Robinson N. Online Biometric Authentication Using Subject-Specific Band Power features of EEG. In: Proceedings of the 2017 International Conference on Cryptography, Security and Privacy - ICCSP ’17 [Internet]. Wuhan, China: ACM Press; 2017 [cited 2022 May 19]. p. 136–41. Available from: http://dl.acm.org/citation.cfm?doid=3058060.3058068
11. Yap HY, Choo YH, Mohd Yusoh ZI, Khoh WH. Person authentication based on eye-closed and visual stimulation using EEG signals. Brain Inform. 2021 Oct 11;8(1):21.
12. Bashar MdK, Chiaki I, Yoshida H. Human identification from brain EEG signals using advanced machine learning method EEG-based biometrics. In: 2016 IEEE EMBS Conference on Biomedical Engineering and Sciences (IECBES). 2016. p. 475–9.
13. Jayarathne I, Cohen M, Amarakeerthi S. Person identification from EEG using various machine learning techniques with inter-hemispheric amplitude ratio. Pappalardo F, editor. PLOS ONE. 2020 Sep 11;15(9):e0238872.
14. Dan Z, Xifeng Z, Qiangang G. An Identification System Based on Portable EEG Acquisition Equipment. In: 2013 Third International Conference on Intelligent System Design and Engineering Applications. 2013. p. 281–4.
15. The Wavelet packet decomposition features applied in EEG based authentication system - ProQuest [Internet]. [cited 2022 May 19]. Available from: https://www.proquest.com/openview/935fd57876b78284c811643a195e4403/1?pq-origsite=gscholar&cbl=4998668
16. Nakamura T, Goverdovsky V, Mandic DP. In-Ear EEG Biometrics for Feasible and Readily Collectable Real-World Person Authentication. IEEE Trans Inf Forensics Secur. 2018 Mar;13(3):648–61.
17. Dai Y, Wang X, Li X, Tan Y. Sparse EEG compressive sensing for web-enabled person identification. Measurement. 2015 Oct 1;74:11–20.
18. Moctezuma LA, Molinas M. EEG-based Subjects Identification based on Biometrics of Imagined Speech using EMD [Internet]. arXiv; 2018 [cited 2022 May 19]. Available from: http://arxiv.org/abs/1809.06697
19. Sooriyaarachchi J, Seneviratne S, Thilakarathna K, Zomaya AY. MusicID: A Brainwave-based User Authentication System for Internet of Things [Internet]. arXiv; 2020 [cited 2022 May 19]. Available from: http://arxiv.org/abs/2006.01751
20. Koike-Akino T, Mahajan R, Marks TK, Wang Y, Watanabe S, Tuzel O, et al. High-accuracy user identification using EEG biometrics. In: 2016 38th Annual International Conference of the IEEE Engineering in Medicine and Biology Society (EMBC). 2016. p. 854–8.
21. Abo-Zahhad M, Ahmed SM, Abbas SN. A Novel Biometric Approach for Human Identification and Verification Using Eye Blinking Signal. IEEE Signal Process Lett. 2015 Jul;22(7):876–80.

22. Gopal SRK, Shukla D. Concealable Biometric-based Continuous User Authentication System An EEG Induced Deep Learning Model. In: 2021 IEEE International Joint Conference on Biometrics (IJCB). 2021. p. 1–8.
23. Moctezuma LA, Torres-García AA, Villaseñor-Pineda L, Carrillo M. Subjects identification using EEG-recorded imagined speech. Expert Syst Appl. 2019 Mar 15;118:201–8.
24. Zhang R, Yan B, Tong L, Shu J, Song X, Zeng Y. Identity Authentication Using Portable Electroencephalography Signals in Resting States. IEEE Access. 2019;7:160671–82.
25. Kaur B, Singh D, Roy PP. A Novel framework of EEG-based user identification by analyzing music-listening behavior. Multimed Tools Appl. 2017 Dec;76(24):25581–602.
26. Lu L, Mao J, Wang W, Ding G, Zhang Z. An EMG-Based Personal Identification Method Using Continuous Wavelet Transform and Convolutional Neural Networks. In: 2019 IEEE Biomedical Circuits and Systems Conference (BioCAS). 2019. p. 1–4.
27. Lu L, Mao J, Wang W, Ding G, Zhang Z. A Study of Personal Recognition Method Based on EMG Signal. IEEE Trans Biomed Circuits Syst. 2020 Aug;14(4):681–91.
28. Raurale SA, McAllister J, Rincón JMD. EMG Biometric Systems Based on Different Wrist-Hand Movements. IEEE Access. 2021;9:12256–66.
29. Cao Y, Zhang Q, Li F, Yang S, Wang Y. PPGPass: Nonintrusive and Secure Mobile Two-Factor Authentication via Wearables. In: IEEE INFOCOM 2020 - IEEE Conference on Computer Communications. 2020. p. 1917–26.
30. Jindal V, Birjandtalab J, Pouyan MB, Nourani M. An adaptive deep learning approach for PPG-based identification. In: 2016 38th Annual International Conference of the IEEE Engineering in Medicine and Biology Society (EMBC). 2016. p. 6401–4.
31. Everson L, Biswas D, Panwar M, Rodopoulos D, Acharyya A, Kim CH, et al. BiometricNet: Deep Learning based Biometric Identification using Wrist-Worn PPG. In: 2018 IEEE International Symposium on Circuits and Systems (ISCAS). 2018. p. 1–5.
32. Zhao T, Wang Y, Liu J, Chen Y, Cheng J, Yu J. TrueHeart: Continuous Authentication on Wrist-worn Wearables Using PPG-based Biometrics. In: IEEE INFOCOM 2020 - IEEE Conference on Computer Communications. 2020. p. 30–9.
33. Khan MU, Aziz S, Zainab A, Tanveer H, Iqtidar K, Waseem A. Biometric System using PCG Signal Analysis: A New Method of Person Identification. In: 2020 International Conference on Electrical, Communication, and Computer Engineering (ICECCE). 2020. p. 1–6.
34. Zhao Z, Shen Q. A human identification system based on Heart sounds and Gaussian Mixture Models. In: 2011 4th International Conference on Biomedical Engineering and Informatics (BMEI). 2011. p. 597–601.
35. Cheng X, Wang P, She C. Biometric Identification Method for Heart Sound Based on Multimodal Multiscale Dispersion Entropy. Entropy. 2020 Feb;22(2):238.
36. Fahad I, Apu MAR, Ghosh A, Fattah SA. Phonocardiogram Heartbeat Segmentation and Autoregressive Modeling for Person Identification. In: TENCON 2019 - 2019 IEEE Region 10 Conference (TENCON). 2019. p. 1942–6.
37. Beritelli F. A multiband approach to human identity verification based on PhonoCardioGram signal analysis. In: 2008 Biometrics Symposium. 2008. p. 71–6.
38. Biometrics from heart sounds: Evaluation of a new approach based on wavelet packet cepstral features using HSCT-11 database | Elsevier Enhanced Reader [Internet]. [cited 2022 May 15]. Available from: https://reader.elsevier.com/reader/sd/pii/S0045790616301057?token=D2A7AA1E4156050E1751CFEB1BC178735B73E96F7E7E30F5C783B69E16AEDA594170AF51FC0CDD2407F6E67DAFA6091F&originRegion=us-east-1&originCreation=20220515052243
39. Kim J, Lee KB, Hong SG. Random forest based-biometric identification using smart shoes. In: 2017 Eleventh International Conference on Sensing Technology (ICST). 2017. p. 1–4.
40. Jeon S, Yoon HJ, Lee YS, Son SH, Eun Y. Poster Abstract: Biometric Gait Identification for Exercise Reward System using Smart Earring. :2.
41. Cola G, Avvenuti M, Musso F, Vecchio A. Gait-based authentication using a wrist-worn device. In: Proceedings of the 13th International Conference on Mobile and Ubiquitous Systems: Computing, Networking and Services [Internet]. Hiroshima Japan: ACM; 2016 [cited 2022 May 15]. p. 208–17. Available from: https://dl.acm.org/doi/10.1145/2994374.2994393
42. Retsinas G, Filntisis PP, Efthymiou N, Theodosis E, Zlatintsi A, Maragos P. Person Identification Using Deep Convolutional Neural Networks on Short-Term Signals from Wearable Sensors. In: ICASSP 2020 - 2020 IEEE International Conference on Acoustics, Speech and Signal Processing (ICASSP). 2020. p. 3657–61.
43. Tao S, Zhang X, Cai H, Lv Z, Hu C, Xie H. Gait based biometric personal authentication by using MEMS inertial sensors. J Ambient Intell Humaniz Comput. 2018 Oct;9(5):1705–12.
44. Moon J, Minaya NH, Le NA, Park HC, Choi SI. Can Ensemble Deep Learning Identify People by Their Gait Using Data Collected from Multi-Modal Sensors in Their Insole? Sensors. 2020 Jan;20(14):4001.
45. Sudhakar SRV, Kayastha N, Sha K. ActID: An efficient framework for activity sensor based user identification. Comput Secur. 2021 Sep 1;108:102319.
46. Ferlini A, Ma D, Harle R, Mascolo C. EarGate: gait-based user identification with in-ear microphones. In: Proceedings of the 27th Annual International Conference on

Mobile Computing and Networking [Internet]. New Orleans Louisiana: ACM; 2021 [cited 2022 May 15]. p. 337–49. Available from: https://dl.acm.org/doi/10.1145/3447993.3483240
47. Musale P, Baek D, Werellagama N, Woo SS, Choi BJ. You Walk, We Authenticate: Lightweight Seamless Authentication Based on Gait in Wearable IoT Systems. IEEE Access. 2019;7:37883–95.
48. Baek D, Musale P, Ryoo J. Walk to Show Your Identity: Gait-based Seamless User Authentication Framework Using Deep Neural Network. In: The 5th ACM Workshop on Wearable Systems and Applications - WearSys '19 [Internet]. Seoul, Republic of Korea: ACM Press; 2019 [cited 2022 May 15]. p. 53–8. Available from: http://dl.acm.org/citation.cfm?doid=3325424.3329666
49. Johnston AH, Weiss GM. Smartwatch-based biometric gait recognition. In: 2015 IEEE 7th International Conference on Biometrics Theory, Applications and Systems (BTAS). 2015. p. 1–6.
50. Al-Naffakh N, Clarke N, Li F. Continuous User Authentication Using Smartwatch Motion Sensor Data. In: Gal-Oz N, Lewis PR, editors. Trust Management XII. Cham: Springer International Publishing; 2018. p. 15–28. (IFIP Advances in Information and Communication Technology).
51. Acar A, Aksu H, Uluagac AS, Akkaya K. A Usable and Robust Continuous Authentication Framework Using Wearables. IEEE Trans Mob Comput. 2021 Jun;20(6):2140–53.
52. Rahman KA, Alam N, Musarrat J, Madarapu A, Hossain MS. Smartwatch Dynamics: A Novel Modality and Solution to Attacks on Cyber-behavioral Biometrics for Continuous Verification? In: 2020 International Symposium on Networks, Computers and Communications (ISNCC). 2020. p. 1–5.
53. Griswold-Steiner I, Matovu R, Serwadda A. Wearables-Driven Freeform Handwriting Authentication. IEEE Trans Biom Behav Identity Sci. 2019 Jul;1(3):152–64.
54. Buriro A, Van Acker R, Crispo B, Mahboob A. AirSign: A Gesture-Based Smartwatch User Authentication. In: 2018 International Carnahan Conference on Security Technology (ICCST). 2018. p. 1–5.
55. Weiss GM, Yoneda K, Hayajneh T. Smartphone and Smartwatch-Based Biometrics Using Activities of Daily Living. IEEE Access. 2019;7:133190–202.
56. Saleheen N, Ullah MA, Chakraborty S, Ones DS, Srivastava M, Kumar S. WristPrint: Characterizing User Re-identification Risks from Wrist-worn Accelerometry Data. In: Proceedings of the 2021 ACM SIGSAC Conference on Computer and Communications Security [Internet]. Virtual Event Republic of Korea: ACM; 2021 [cited 2022 Apr 20]. p. 2807–23. Available from: https://dl.acm.org/doi/10.1145/3460120.3484799
57. Lee S, Choi W, Lee DH. Usable User Authentication on a Smartwatch using Vibration. In: Proceedings of the 2021 ACM SIGSAC Conference on Computer and Communications Security [Internet]. New York, NY, USA: Association for Computing Machinery; 2021 [cited 2022 May 14]. p. 304–19. (CCS '21). Available from: https://doi.org/10.1145/3460120.3484553
58. Jo YC, Kim HN, Hong HK, Choi YS, Jung SW, Kang JH, et al. Feasibility of a wearable band type biometrie devices using skin spectroscopy. In: 2017 IEEE International Conference on Systems, Man, and Cybernetics (SMC). 2017. p. 1903–7.
59. Maiorana E, Massaroni C. Biometric Recognition based on Heart-Induced Chest Vibrations. In: 2021 IEEE International Workshop on Biometrics and Forensics (IWBF). 2021. p. 1–6.
60. Hsu PY, Hsu PH, Liu HL. Exploring Seismocardiogram Biometrics with Wavelet Transform. In: 2020 25th International Conference on Pattern Recognition (ICPR). 2021. p. 4450–7.
61. Takeda T, Kuramoto K, Kobashi S, Hata Y. Biometrics Personal Identification by Wearable Pressure Sensor. In: 2012 Fifth International Conference on Emerging Trends in Engineering and Technology. 2012. p. 120–3.
62. Chauhan J, Hu Y, Seneviratne S, Misra A, Seneviratne A, Lee Y. BreathPrint: Breathing Acoustics-based User Authentication. In: Proceedings of the 15th Annual International Conference on Mobile Systems, Applications, and Services [Internet]. New York, NY, USA: Association for Computing Machinery; 2017 [cited 2022 May 20]. p. 278–91. (MobiSys '17). Available from: https://doi.org/10.1145/3081333.3081355
63. Chauhan J, Rajasegaran J, Seneviratne S, Misra A, Seneviratne A, Lee Y. Performance Characterization of Deep Learning Models for Breathing-based Authentication on Resource-Constrained Devices. Proc ACM Interact Mob Wearable Ubiquitous Technol. 2018 Dec 27;2(4):158:1-158:24.
64. Raji RK, Adjeisah M, Miao X, Wan A. A novel respiration pattern biometric prediction system based on artificial neural network. Sens Rev. 2020;40(1):8–16.
65. Gao Y, Wang W, Phoha VV, Sun W, Jin Z. EarEcho: Using Ear Canal Echo for Wearable Authentication. Proc ACM Interact Mob Wearable Ubiquitous Technol. 2019 Sep 9;3(3):81:1-81:24.
66. Lei H, Liu J, Zou Y, Wu K. Smart earpieces that know who you are quietly: poster abstract. In: Proceedings of the 18th Conference on Embedded Networked Sensor Systems [Internet]. New York, NY, USA: Association for Computing Machinery; 2020 [cited 2022 May 20]. p. 721–2. Available from: https://doi.org/10.1145/3384419.3431254
67. Noh HW, Sim JY, Ahn CG, Ku Y. Electrical Impedance of Upper Limb Enables Robust Wearable Identity Recognition against Variation in Finger Placement and

Environmental Factors. Biosensors. 2021 Oct;11(10):398.
68. Schneegass S, Oualil Y, Bulling A. SkullConduct: Biometric User Identification on Eyewear Computers Using Bone Conduction Through the Skull. In: Proceedings of the 2016 CHI Conference on Human Factors in Computing Systems [Internet]. New York, NY, USA: Association for Computing Machinery; 2016 [cited 2022 May 20]. p. 1379–84. (CHI '16). Available from: https://doi.org/10.1145/2858036.2858152
69. Biometric recognition using wearable devices in real-life settings | Elsevier Enhanced Reader [Internet]. [cited 2022 May 20]. Available from: https://reader.elsevier.com/reader/sd/pii/S0167865521001070?token=31A9A540ED7CE38DCC52112B654DD6F5E99F92E2FD11F60A49C55B6AA8F82B9DFEE57C4C521ACDCCC9187B2E5E18A446&originRegion=us-east-1&originCreation=20220520180511
70. Ekiz D, Can YS, Dardagan YC, Ersoy C. Can a Smartband be Used for Continuous Implicit Authentication in Real Life. IEEE Access. 2020;8:59402–11.
71. Vhaduri S, Poellabauer C. Multi-Modal Biometric-Based Implicit Authentication of Wearable Device Users. IEEE Trans Inf Forensics Secur. 2019 Dec;14(12):3116–25.
72. Vhaduri S, Poellabauer C. Wearable device user authentication using physiological and behavioral metrics. In: 2017 IEEE 28th Annual International Symposium on Personal, Indoor, and Mobile Radio Communications (PIMRC). 2017. p. 1–6.
